# Epidemiological Aspects of Zoonotic Cutaneous Leishmaniasis (ZCL) in Iran, the Middle East, and Worldwide: A Comprehensive Systematic Review

**DOI:** 10.1101/2024.05.06.24306918

**Authors:** Hedayatullah jamali, Saied Bokaie

## Abstract

**Introduction and objectives:** Zoonotic cutaneous leishmaniasis (ZCL) remains a significant health problem, particularly in Iran, where 15.8 cases per 100,000 people were reported in 2019. Iran accounted for more than half of all new cases in the Eastern Mediterranean region in 2008. entified several countries in the Middle East and North Africa, including Syria, Afghanistan, Iran, Pakistan, Iraq, Yemen, Tunisia, Morocco, Libya and Egypt, as having the highest rates of this disease, which is a major burden in the rural areas of these regions. ZCL is a major global health problem, especially in countries such as Iran, the Middle East and North Africa.ZCL is a major global health problem, particularly in countries such as Iran, the Middle East and North Africa. Despite the existing research on cutaneous leishmaniasis, few studies focus exclusively on the epidemiologic aspects of ZCL. This systematic study aims to fill this gap by investigating the geographical distribution and cause of occurrence and identifying reservoirs, vectors and sites of ZCL occurrence, focusing on high-risk areas such as Algeria and Tunisia in North Africa in figure 8. Areas suitable for vector-borne transmission and ZCL reservoirs are expected to increase in the future. Understanding the ecological links between vectors, reservoirs and the Leishmania parasite is crucial for the development of effective control and prevention strategies. It is expected that the regions favorable for vector-borne transmission and the reservoirs of zoonotic cutaneous leishmaniasis (ZCL) will expand as environmental conditions evolve. To develop effective control and prevention strategies, it is important to understand the ecological interplay between vectors, reservoirs and the leishmaniasis parasite. Climate change is expected to exacerbate the threat of ZCL by 2050, potentially increasing the suitability of habitats for vectors and reservoirs. This study uniquely examines the epidemiologic aspects of ZCL globally, filling a gap in the current literature that predominantly addresses cutaneous leishmaniasis in a broader context.

**Results:** This study provides a detailed insight into the increasing prevalence of zoonotic cutaneous leishmaniasis (ZCL) in Iran. It identifies Iran, where 18 out of 31 provinces are affected, as a critical area, especially the central provinces. Key factors such as Leishmania reservoirs in rodents, emerging reservoirs, and specific vectors contribute significantly to the transmission of the disease and are influenced by environmental and climatic conditions in Iran, the Middle East, and North Africa. The study highlights new hotspots in Iran, such as Beyza district in Fars province and regions in Isfahan and Razavi Khorasan provinces, which indicate the dynamic nature of the spread of ZCL associated with urbanization and climate change. Predictive modeling suggests that an increase in ZCL may soon occur in northwestern Iran due to suitable environmental conditions for the vectors and reservoirs. On the other hand, the identification of new reservoirs was an important result. These included different hedgehog species (Paraechinus aethiopicus, Atelerix algirus, and Hemiechinus auritus) in Algeria, Tunisia, and Iran as well as calomyscid rodents in Shiraz, the capital of the Iranian province of Fars. Interestingly, specific vectors and reservoirs make Tunisia and Algeria high-risk areas in North Africa, emphasizing the need for regional integrated control measures. The results highlight a crucial gap in the research and control of zoonotic cutaneous leishmaniasis outside Iran. In the last ten years, the focus has been less on the Middle East and Central Asia, although the disease is widespread in these regions.

**Conclusion:** This review emphasizes the critical need for a combined approach to the control and prevention of zoonotic cutaneous leishmaniasis (ZCL). This study emphasizes the role of climate change and urban expansion in influencing disease dynamics. The identification of new endemic areas and prediction of future hotspots in Iran will provide valuable insights for targeting prevention and control measures. This underlines the importance of regional collaboration and adaptive strategies in the Middle East and North Africa (MENA) countries to effectively tackle this neglected tropical disease as part of the One Health approach. As the first comprehensive study on the epidemiology of ZCL, this study fills a significant gap in the literature and provides a foundation for future research and public health interventions to mitigate the global impact of ZCL. This systematic review highlights the complicated and multifaceted nature of the disease, which is influenced by various vectors, reservoir hosts, and environmental factors. This comprehensive review not only deepens our understanding of the epidemiology of ZCL, but also provides crucial insights for health managers and policy makers. This knowledge can help them to identify high-risk areas, implement targeted prevention measures, and develop effective control programs to combat this endemic disease.

## Introduction

Zoonotic cutaneous leishmaniasis (ZCL) is a serious disease caused by parasites that are transmitted from infected animals to humans through the bite of sandflies. This disease is a major public health problem affecting millions of people in Iran and worldwide. Close contact with infected animals increases the risk of humans becoming infected with the Leishmania parasite. ZCL is not only a health problem, but also represents a significant socio-economic burden, especially in areas where the disease is widespread. In order to develop effective prevention and control strategies, it is important to understand how this disease spreads. Different Leishmania parasite species can be transmitted by infected female sandflies and cause a range of symptoms in humans, from self-healing skin lesions to potentially fatal internal infections, depending on which parasite is involved. (1–4).

Zoonotic cutaneous leishmaniasis (ZCL) is a serious disease transmitted by insects. It affects approximately 12 million people in 98 countries where it is widespread. As shown in Figures 1 and 4, there are nearly 2 million new cases each year. The complex nature of this disease, including its various animal hosts and insect vectors, underscores the public health importance of this disease (5–9,1).

**Figure 1.**
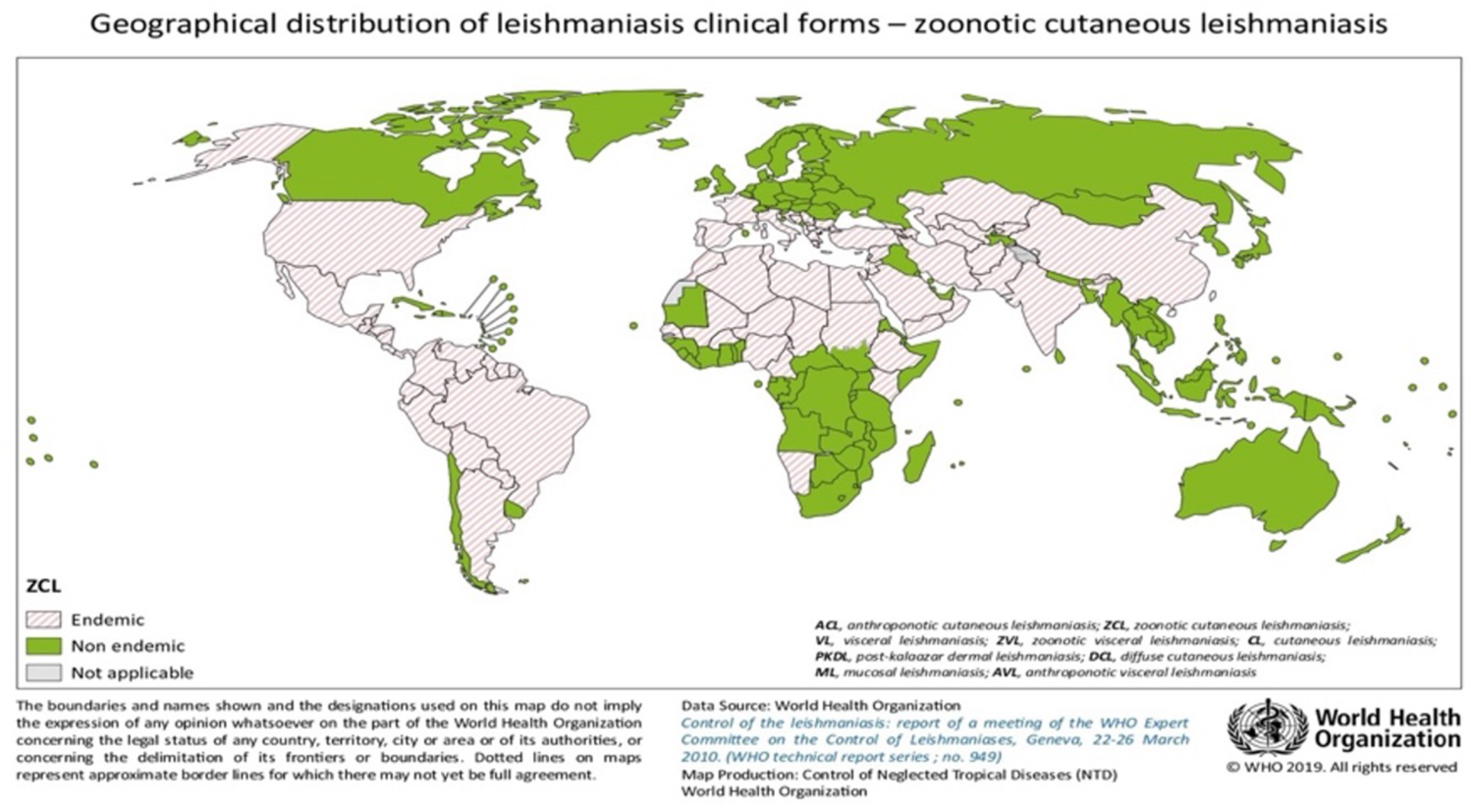
Geographical distribution of clinical forms of ZCL in the countries of the world

Figure 1 shows the geographical distribution of clinical forms of ZCL in countries around the world^[1]^.

Figure 1 shows the geographical distribution of zoonotic cutaneous leishmaniasis (ZCL). Dark green colored areas are not endemic, while the diagonally striped areas are endemic for ZCL. The countries of the Middle East and North Africa such as Iran, Iraq, Syria, Saudi Arabia, Yemen, Libya, Tunisia, Algeria, and Morocco are designated as endemic regions for ZCL. In addition, ZCL also occurs in parts of sub-Saharan Africa, Central Asia, and South America.

Zoonotic cutaneous leishmaniasis (ZCL) is a skin disease that can cause disfiguring lesions and permanent scarring if left untreated. This can lead to serious social stigmatization and disability for those affected. The spread of ZCL is determined by complex interactions between the environment, human behavior and the dynamics between disease-transmitting insects, animal hosts and humans. In regions where ZCL is widespread, such as Iran and throughout the Middle East, it continues to pose challenges to the healthcare system. A closer look at the epidemiology of the disease is needed. ZCL has a complex transmission cycle involving animal reservoirs such as rodents and other small mammals (10–15).

Zoonotic cutaneous leishmaniasis (ZCL) is a disease caused by a tiny parasite called Leishmania major. This parasite is transmitted to humans by wild rodents such as Psammomys obesus through the bite of sand flies. People living near these rodents and sand flies can also become infected. The number of ZCL cases in humans depends on the environment and climate. Heavy rainfall can lead to more chenopods, a plant species that eats Psammomys obesus. When there are more Psammomys obesus, the parasites they carry can more easily spread to the sand flies that bite them. This means that humans are more likely to be infected in the future. These results show that ZCL is strongly influenced by climatic factors (16).

The transmission of this disease depends on environmental and societal factors. Therefore, a comprehensive approach is needed to understand the risk and extent of transmission (17). Leishmaniasis is increasing in many countries, including Iran, due to climate change, refugee flows and urbanization (18(. The control of leishmaniasis is a major challenge. In recent years, the disease has been on the rise in different parts of the country and worldwide. Health authorities and armed forces deployed in border areas need to have comprehensive information on clinical symptoms, control methods, prevention, diagnosis and treatment. With careful and necessary planning, they can prevent the spread of this disease (19).

Maintaining strong disease surveillance systems and effective pest control measures is critical to preventing infection. Awareness campaigns are also important to improve understanding of the disease (20). Cutaneous leishmaniasis is a disease that is prevalent worldwide. About one third of cases occur in each of the three main regions: North and South America, the Mediterranean region and Western Asia from the Middle East to Central Asia. The 10 countries with the most reported cases are Afghanistan, Algeria, Colombia, Brazil, Iran, Syria, Ethiopia, North Sudan, Costa Rica and Peru. Together, these countries account for 70–75% of global cases of cutaneous leishmaniasis. Areas where the zoonotic form of cutaneous leishmaniasis (caused by the parasite Leishmania major) occurs include Afghanistan, Egypt, Iran, Iraq, Jordan, Libya, Morocco, Palestine, Pakistan, Saudi Arabia, Sudan, Syria, Tunisia and Yemen (21–22).

Figures 1-3 update these statistics and include many more countries. Zoonotic cutaneous leishmaniasis (ZCL) is a widespread disease that occurs in certain regions. It is caused by the parasite Leishmania major and is spread by the sand fly Phlebotomus papatasi. This disease mainly affects disadvantaged communities in Iran, the Middle East and other parts of the world. The changing patterns of this disease require a thorough investigation to understand its current status and future evolution.

Figure 2 shows the geographical distribution of Leishmania major detected in different parts of the world. (Source^[1]^: World Health Organization website)

**Figure 2.**
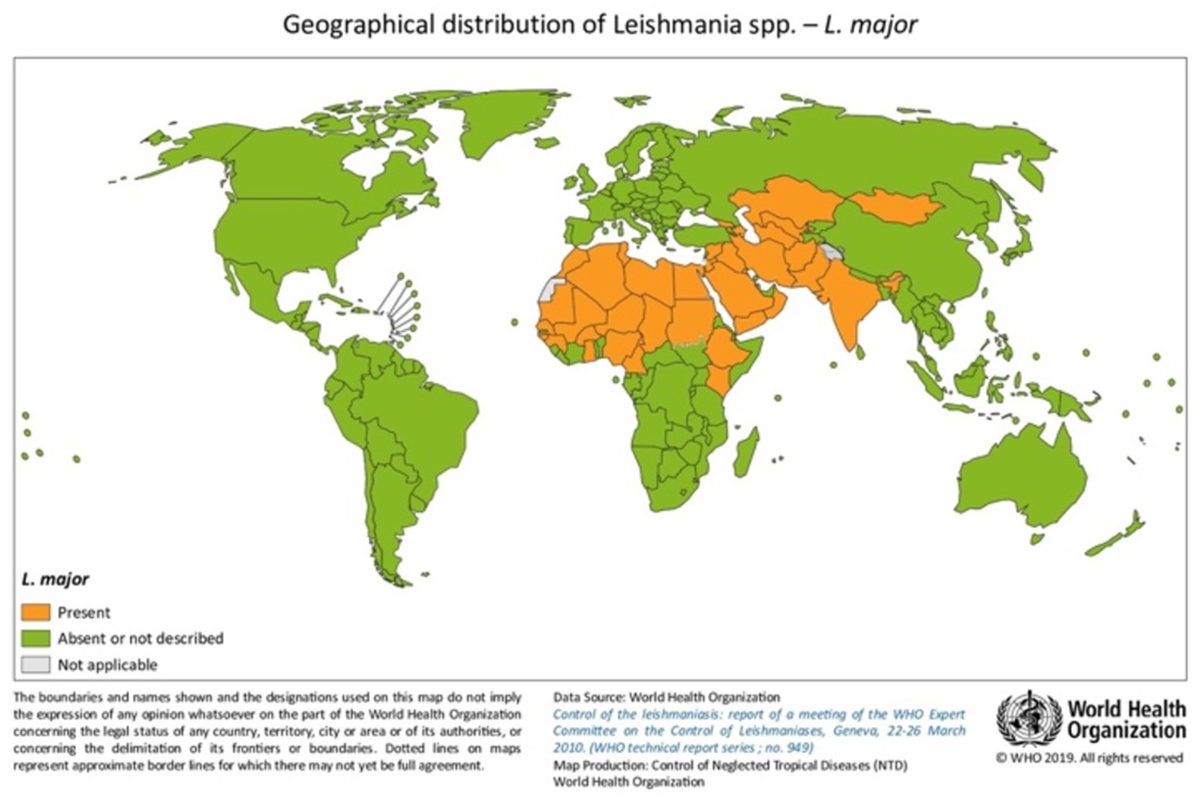
The geographical distribution of Leishmania major is confirmed in different parts of the world.

**Figure 3.**
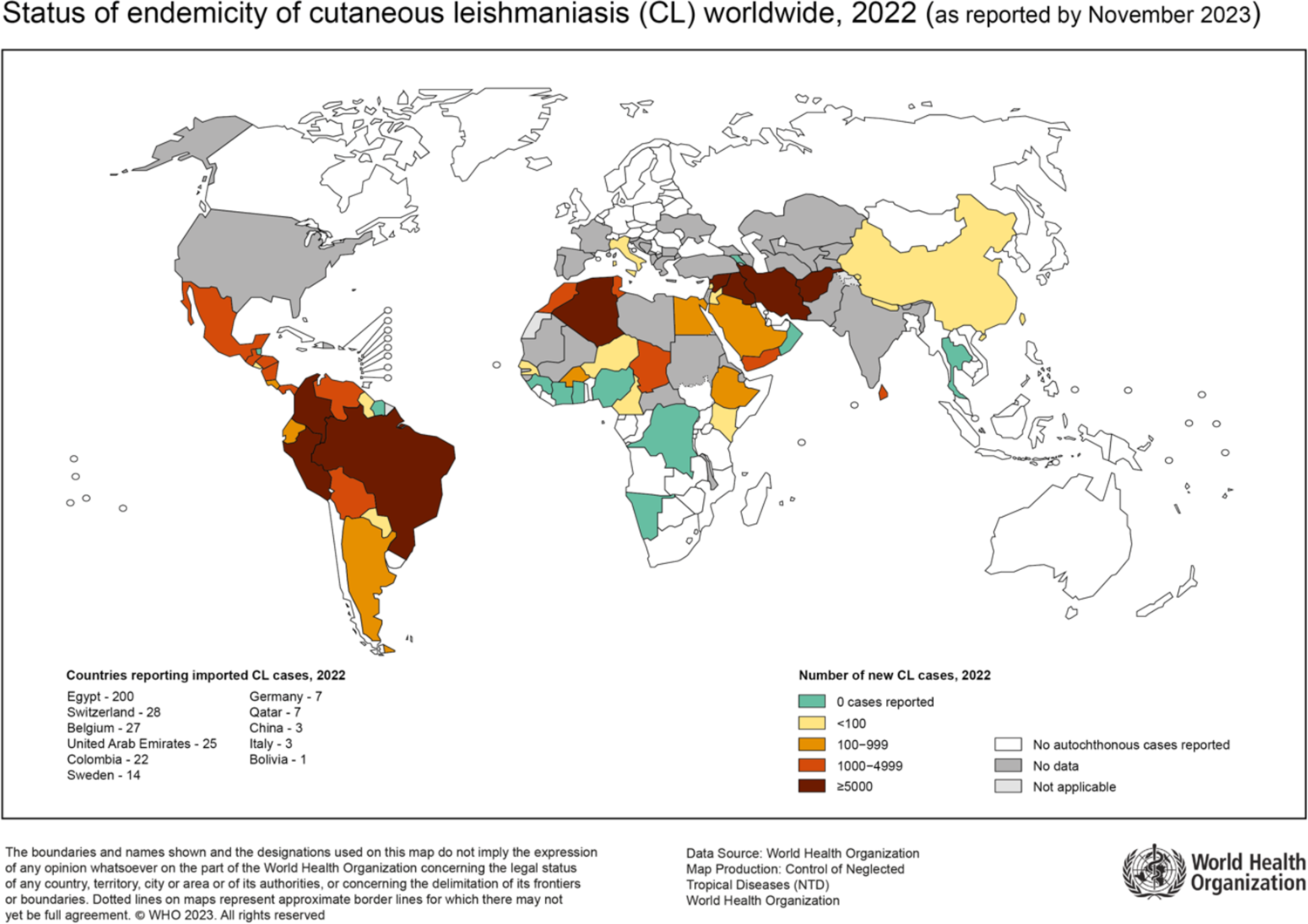
The endemic situation of cutaneous leishmaniasis in 2022 in the countries of the world

Figure 2 also shows that the parasite Leishmania major is mainly found in the Middle East, North Africa and parts of East Africa. In contrast, it is largely absent or not well documented in Europe, North America and most of Asia. The presence and distribution of L. major in South America varies, with some countries reporting its existence and others not.

Figure 3 shows the endemic situation of cutaneous leishmaniasis in 2022 in various countries around the world ^[2]^.

Leishmaniasis is a widespread disease that occurs both in the New World (South and Central America, Mexico) and in the Old World (Europe, Africa, Central Asia and the Indian subcontinent). There are three main forms of the disease: the skin, the mucous membranes and the viscera. Although cutaneous leishmaniasis is not fatal, it can cause severe skin lesions and psychological problems. Over 90% of cases occur in certain regions, including Afghanistan, Pakistan, Iran, Iraq, Syria, Jordan, Algeria, Tunisia, Morocco, Saudi Arabia (Old World), Brazil and Peru (New World). About 12 million people in 98 countries are infected with leishmaniasis, and about 1 million new cases are diagnosed each year, resulting in 50,000–100,000 deaths (12).

The complex relationship between ZCL disease transmission and changing risk environments needs to be studied in more detail. The aim is to understand how the interactions between biological, environmental and social factors drive the incidence of the disease in local or regional areas. Although the epidemiology of ZCL has been well characterised in many established endemic zones, there are still gaps in predicting changing spatial patterns, identifying locations at risk of new endemic establishment, and quantifying infection levels. This information is crucial for setting control priorities in the different ecosystems harboring potential vector or animal reservoir species. In our view, joint programs that take a holistic approach have a better chance of reducing the burden of ZCL in North Africa and the Middle East. This approach should include management of reservoir hosts and sandflies as well as consideration of human behavior and environmental needs as part of the One Health approach (22–25).

Over the past 10 years, approximately 1.5 million new cases of zoonotic cutaneous leishmaniasis (ZCL) have been reported each year. ZCL occurs in over 80 countries, with high rates in parts of the Middle East, Africa and South America. The countries with the most ZCL cases are Brazil, Algeria and Iran, as the ecosystems favor the spread of the disease (1(. Areas with high ZCL tend to have more health problems due to factors such as poor infrastructure, poverty, lack of access to healthcare and limited resources for pest control. In regions with lower ZCL, sporadic cases often occur in travelers returning from endemic areas or imported cases from animal populations. ZCL is a significant public health problem in North Africa, the Middle East and parts of Central/South Asia, with Iran being one of the most severely affected countries (16,26–27).

In 2015, the World Health Organization (WHO) reported on the incidence of zoonotic cutaneous leishmaniasis (ZCL) in North Africa and West Asia and on the incidence of cutaneous leishmaniasis in North Africa and the Middle East. These results are shown in Figures 4 and 5 respectively.

**Figure 4.**
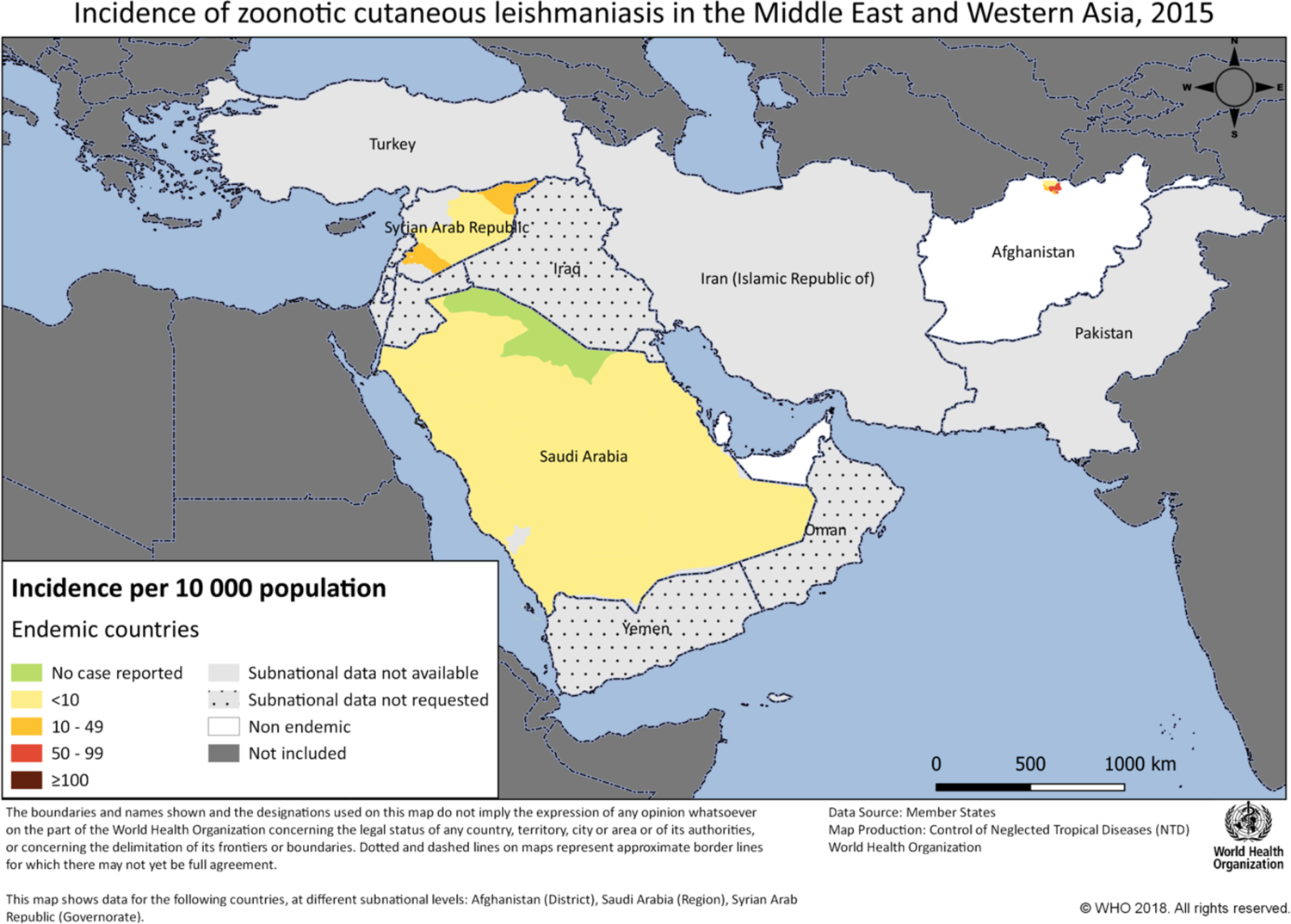
Incidence of zoonotic cutaneous leishmaniasis (ZCL) in North Africa and Western Asia,2015

**Figure 5.**
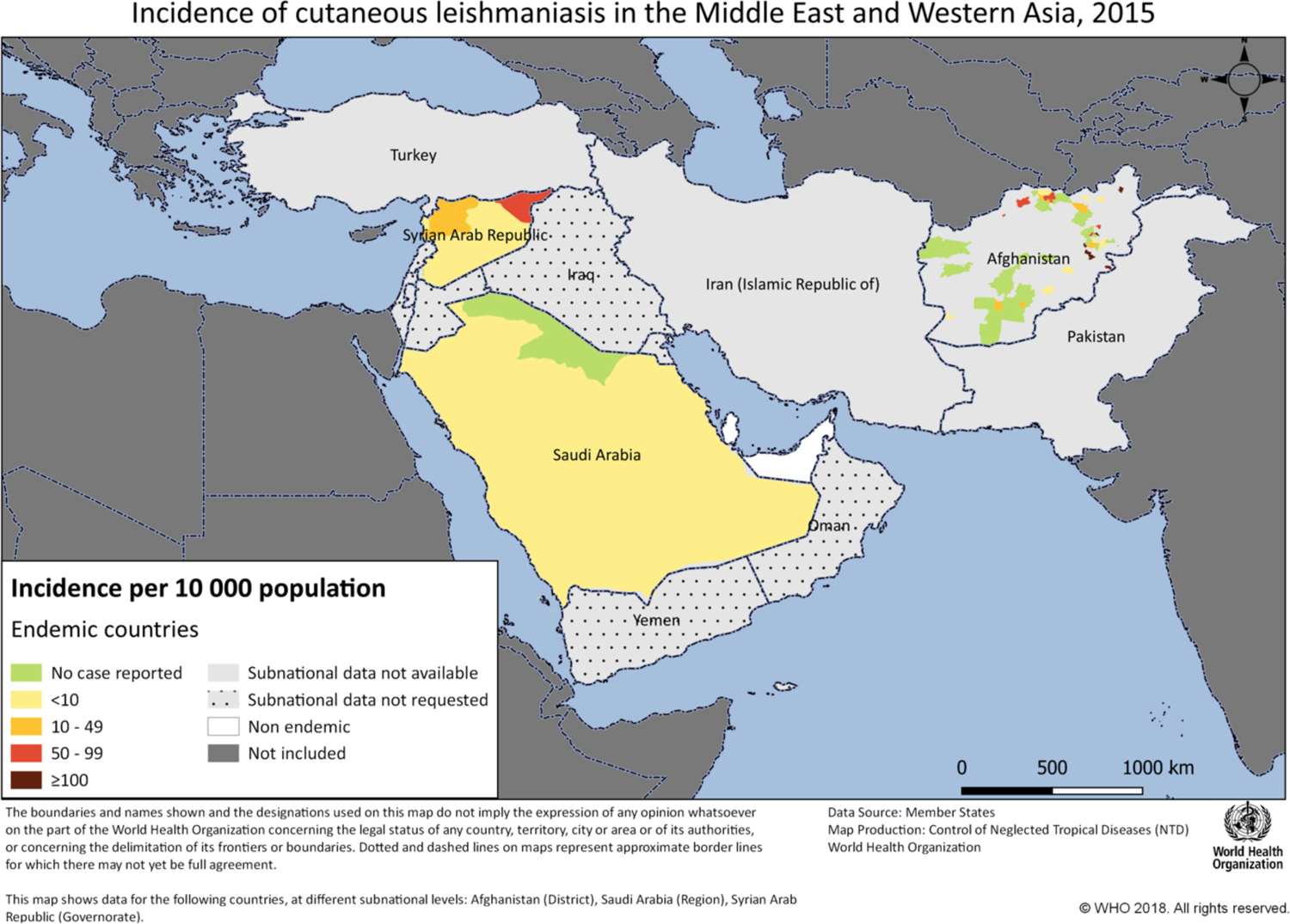
Incidence of Cutaneous Leishmaniasis in North Africa and the Middle East,2015 (Figures 4 **and 5****, quoted by WHO^[1]^,^[2]^)** Source:https://leishinfowho-cc55.es/wp-content/uploads/2022/07/pdfs/essential-maps/regional-maps/North_Africa_Middle_east_CL.png https://leishinfowho-cc55.es/wp-content/uploads/2022/07/pdfs/essential-maps/regional-maps/middle_east_ZCL.png: Source^2^

In Yurche¬nko et al. (2022), the researchers examined historical and current data on Leishmania species. They collected combined infection data from several countries, including Turkmenistan, Uzbekistan, Kazakhstan, Kyrgyzstan, Iran, China and Mongolia. The researchers focused on a group of closely related Leishmania species (L. major, L. turanica and L. gerbilli) that use similar animal reservoirs and vectors. In a separate study conducted in Pakdasht, a city in the Iranian province of Tehran, the Leishmania species diagnosed in all patients was found to be L. major. This suggests that Pakdasht may be a new region where an outbreak of zoonotic cutaneous leishmaniasis is occurring. Therefore, it is crucial to identify the prevalence and new areas affected by this disease in different parts of Iran. In addition, new outbreaks of this disease have been reported in the cities of Jahrom and Darab in Fars province in southwestern Iran (28–31).

Cutaneous leishmaniasis is a major public health problem in many warm regions, including Iran. In 2008, more than half (54%) of new cases of zoonotic cutaneous leishmaniasis (ZCL) in eastern Mediterranean countries were reported from Iran. This parasitic infection poses a challenge in tropical and subtropical regions.

Zoonotic cutaneous leishmaniasis (ZCL) is an important public health problem, particularly in Iran, and reflects a larger global problem. The different forms of ZCL and the genetic diversity of Leishmania parasites result in different clinical symptoms, highlighting the complexity of this disease (36–37). This review aims to answer the following key questions: What are the main disease vectors, the main animal hosts and the factors influencing the incidence (incidence or prevalence) and spread (transmission) of zoonotic cutaneous leishmaniasis (ZCL)? What are the current epidemiologic trends of zoonotic cutaneous leishmaniasis in Iran, the Middle East and worldwide, and what factors influence its spread and control?

Iran is among the 10 countries with the highest number of cases of cutaneous leishmaniasis (CL). The main causes are Leishmania major and L. tropica, which lead to zoonotic CL (ZCL) and anthroponotic CL (ACL), respectively (39–40). ZCL is a major health problem worldwide, especially in the Middle East, where environmental and political factors favour its spread. In Iran, a high-risk country, there are many cases of ZCL, mostly in rural and suburban areas. Agricultural workers are often the most affected as they are more exposed to the sand fly vectors. The geographic spread of ZCL outbreaks is also favoured by seasonal migrations of the human population and reservoir hosts, which complicates surveillance (41–42).

Leishmaniasis is a serious disease that affects over 350 million people in 98 countries. Every year, around 0.7–1.2 million new cases of this skin disease are reported worldwide. Iran is one of the countries severely affected by this disease. There are two main types - zoonotic cutaneous leishmaniasis (ZCL) and anthroponous cutaneous leishmaniasis (ACL) - which are endemic in different regions. Understanding the specific type of cutaneous leishmaniasis (CL) is crucial for the development of effective prevention strategies. According to the World Health Organization, the main cause of ZCL in Iran is the parasite Leishmania major, while L. tropica is the main cause of ACL. In addition, Iran has also been severely affected by the COVID-19 pandemic, which has exacerbated the country’s public health challenges. To address these issues, a clear understanding of the epidemiology and transmission patterns of leishmaniasis in Iran is crucial for implementing targeted control measures and mitigating the overall impact on the population (43–44,104).

Phlebotomus mosquitoes are common vectors in the Philippines. Cases of cutaneous leishmaniasis (CL) have been reported from areas such as the provinces of Isfahan, Fars, Khorasan, Khuzestan and Kerman in Iran. Other known CL hotspots include parts of Isfahan, south of Fars, Razavi-North-South Khorasan, Mazandaran, Khuzestan, Ilam, Bushehr, Hormozgan, Sarkhs, Kashmar, Lotfabad, Kashan and Damghan. Zoonotic cutaneous leishmaniasis is widespread in rural areas. Epidemiological changes, such as new endemic areas and the spread of leishmaniasis to new regions, have been documented in various studies. The large cities in Iran, including the capital Tehran as well as Isfahan, Shiraz, Mashhad, Tabriz and Ahvaz, have expanded considerably due to migration from the villages and have grown in population. The growth of the cities and the influx of more people into these areas has made it necessary to investigate the spread of diseases such as leishmaniasis in these regions. Eighteen out of 31 provinces in Iran have areas where these diseases are common, and about 80% of the cases are due to zoonotic cutaneous leishmaniasis (ZCL). The remaining percentage is caused by anthroponotic cutaneous leishmaniasis (ACL) (12,45–57).

Iran is one of the most important areas in Asia where cutaneous leishmaniasis (CL) is widespread. Recently, the number of CL cases has increased and the disease has spread to new areas. As there are various risk factors, controlling the disease and implementing effective public health strategies should be a high priority. This will help to treat existing cases and prevent further spread of CL to new areas (48–49). Figure 6 shows the distribution of different types of Leishmania parasites (ZCL and ACL) in different regions of Iran as determined by molecular analysis. This information is from a research article by Hajjaran et al. (2021) (104).

**Figure 6.**
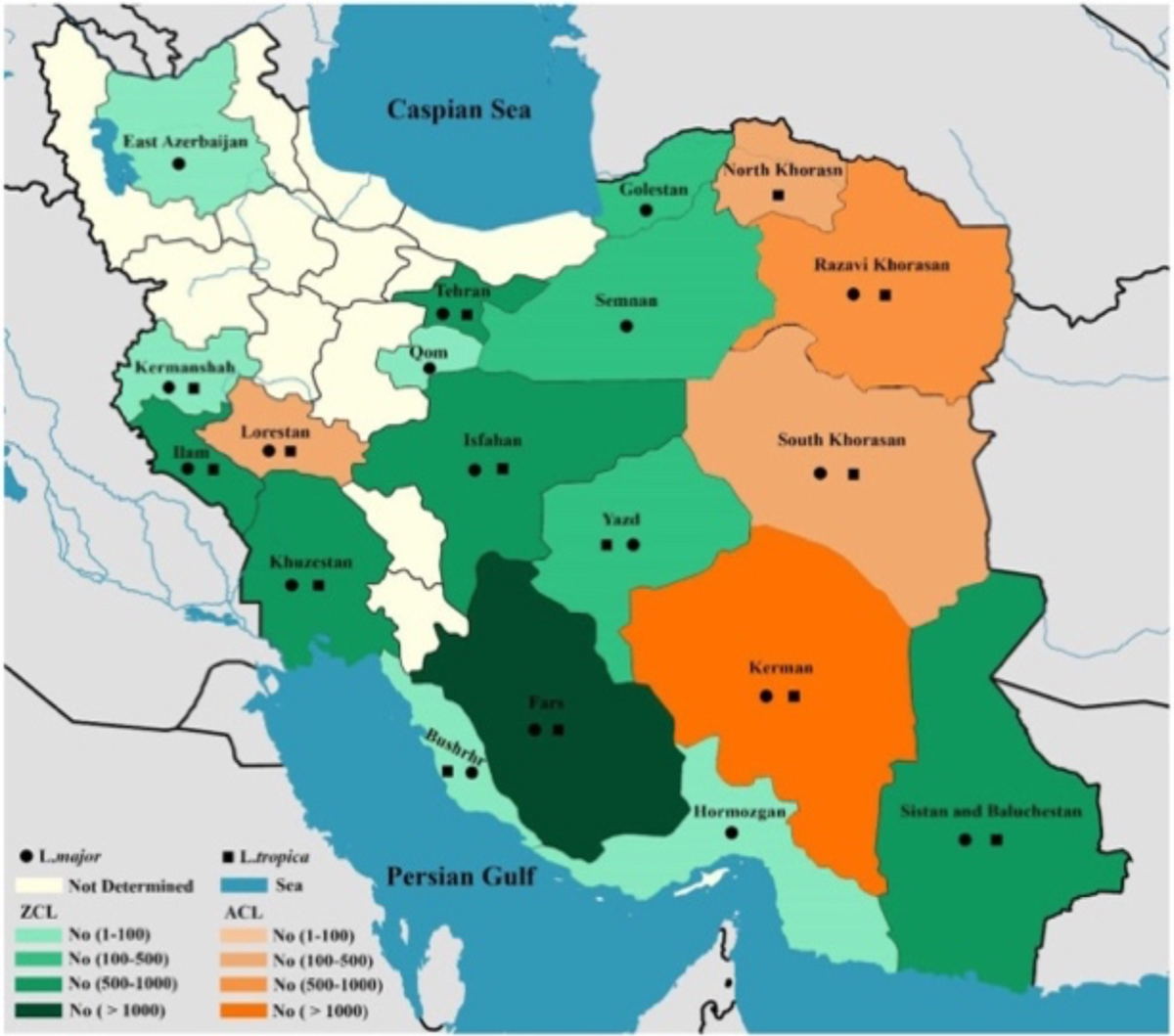
Distribution map of ZCL and ACL Leishmania species determined using molecular methods in different geographical regions of Iran(Source: Article by Hajjaran et al. (2021); with permission).

Zoonotic cutaneous leishmaniasis (ZCL) is a serious health problem in many rural areas of Iran. Wild rats are an important reservoir for this vector-borne disease. An analysis of the locations of ZCL outbreaks shows that 97.8% of cases occur at altitudes below 725 m in arid and semi-arid climates with poor living conditions. Identifying these high-risk areas is crucial to closely monitor and control the spread of ZCL through targeted measures and the provision of resources (49,27).

The fight against this disease has always been part of Iran’s national plans. Despite efforts and investments at national and international levels, the disease has not been brought under control. Instead, new outbreaks have occurred and the disease has continued to spread throughout the country. To choose the right approach to control this disease and improve the success of control programs, we need to understand the epidemiological characteristics of the disease. Zoonotic cutaneous leishmaniasis (ZCL) is widespread in many parts of Iran. The main animal hosts of the L. major parasite in rural Jahrom are Tatera indica and Me. persicus. In the urban areas of Jahrom region, Mu. musculus and Ra. rattus also play a small but notable role in the persistence of the disease (7,30).

Zoonotic cutaneous leishmaniasis (ZCL) is a worrying health problem around the world, particularly in the Middle East. While previous studies have attempted to predict the spread of ZCL based on different environmental factors, the complex interactions between disease vectors (such as the sand fly Phlebotomus papatasi), animal hosts, humans and the environment can influence the spread of the disease. Understanding these interlinked dynamics is crucial for the control of this neglected tropical disease (11). Zoonotic cutaneous leishmaniasis (ZCL) is a widespread disease in Iran caused by the parasite Leishmania major. Close contact with infected animals increases the likelihood of the parasite spreading to humans. Four gerbil species are the main hosts of the ZCL reservoir in different areas of Iran. Rhombomys opimus, Meriones libycus, Meriones hurricane and Tatera indica gerbils. Significant infection was also found in Nesokia indica. This study predicted where these ZCL reservoir host species are found in Iran. The study found that climate and geography are important factors in determining the habitat of these gerbils. In light of climate change, the models used in this study need to be regularly updated with new data. This information should then be used in programs to monitor and track the spread of this disease (3).

Rodents are the main vectors of zoonotic cutaneous leishmaniasis (ZCL). Studies on Leishmania infections in rodents began in 1953 in northeastern Iran. These studies showed that giant gerbils (Rhombomys opimus), Libyan gerbils (Meriones libycus) and Indian gerbils (Tatera indica) play an important role in the transmission of the disease. Previous studies have shown that Leishmania major, L. gerbilli and L. turanica are present in natural populations of gerbils. However, only L. major is pathogenic to humans and responsible for the common process of ZCL development. This emphasises the close relationship between humans and these animals in the transmission of the disease (50,51). Table 4 shows the degree of ZCL risk depending on the reservoir and density of S and their relationship (5)

This table from the study by Ahmed Karmaoui et al. (2022) appears to classify the risk levels of ZCL (zoonotic cutaneous leishmaniasis) based on the presence and combination of certain factors, such as the type of disease reservoir and the presence of a vector. The “+” signs indicate the factors associated with each risk level. For example, the presence of Meriones shawi, Meriones libycus or Phlebotomus papatasi alone is associated with risk level 1. However, as the complexity of the combinations increases, including both reservoirs and disease vectors, the risk increases, with the most complex combination associated with the highest risk level 4. The toolbox in the Arc-GIS software maps risks posed by one or more species that act as reservoirs or vectors. It combines isolated risks with the help of the raster calculator. This produces a global map with risk levels from 1 (very low) to 4 (high).

The geographical distribution of zoonotic cutaneous leishmaniasis (ZCL) has been steadily expanding in Iran recently. Beyza district in Fars province is one of the newly identified endemic areas for ZCL in southern Iran. In addition, a study in Isfahan province has revealed that Aran and Bidgol have emerged as new foci of cutaneous leishmaniasis in humans and animals in central Iran, emphasizing the need to prevent the spread of the disease and control rodents and mosquitoes. Various rodents serve as the main reservoir for zoonotic cutaneous leishmaniasis (ZCL) throughout the Middle East and Central Asia. These findings suggest that uncontrolled and scattered prevention efforts may result in rodents migrating towards human population centers and triggering new waves of infection (23,53).

The spread of zoonotic cutaneous leishmaniasis (ZCL) depends on the presence, number and distribution of the main rodents harboring the Leishmania parasite. The development of these rodents is strongly influenced by environmental and climatic factors. Data show that reported ZCL cases peak seasonally from August to January. The study in Tunisia revealed that rodent density, average temperature, total rainfall and average humidity contribute to the maintenance and increase of ZCL cases with a certain time lag. The gerbil Rhombomys opimus is the primary reservoir host for zoonotic cutaneous leishmaniasis (ZCL) in Iran and neighboring countries (54,39(.) Zoonotic cutaneous leishmaniasis (ZCL) is widespread in various regions of Iran, affecting both rural and urban areas in 18 provinces, including Fars, Isfahan, Kerman, Khuzestan and Lorestan. The disease typically peaks in late summer and fall, which coincides with increased sand fly activity. This suggests that control measures should be implemented before this time to reduce the risk of transmission. Socio-economic factors such as poverty, population displacement and poor infrastructure contribute significantly to ZCL transmission. Environmental and climatic factors such as temperature, humidity and land use changes due to deforestation and irrigation also influence the habitats and behaviour of vectors. Human activities, including construction and population movements, have been associated with the spread of ZCL in Iran (55–57,41,11).

The transfer of ZCL depends on ecological and economic conditions. A multi-layered approach is needed to understand the risk and extent of transmission. This will help to identify key risk factors and understand how different factors interact to influence the spread of the disease at local and regional levels. The occurrence of ZCL is associated with human activities, environmental changes or a combination of these factors. Historical analyses provide valuable insights into ZCL, particularly in Iran, by tracing the evolution of research on this disease since the 19th century (17,58).

This introduction captures the various aspects of ZCL that can be observed in different regions and eras. This forms the basis for a thorough examination of the topic. The aim of this comprehensive overview is not only to discuss the epidemiology of the disease, but also to examine its impact on society and identify ways to control and prevent the disease. Iran is a focus country for this study due to the high number of ZCL cases reported annually. This study investigates the patterns of disease vectors, reservoirs and risk factors associated with the occurrence or prevalence of zoonotic cutaneous leishmaniasis (ZCL). This will help to identify gaps in existing research and make recommendations for future studies and public health interventions.

Zoonotic cutaneous leishmaniasis (ZCL) is a serious public health problem that varies in severity in different regions, including the Middle East and worldwide. Iran, in particular, faces a growing threat due to the diverse environmental conditions that favour the disease-causing organisms and their hosts. ZCL, a major neglected disease, primarily affects economically disadvantaged populations, especially in developing countries. This disease of poverty is a health problem and a major obstacle to local development and prosperity, particularly in indigenous areas. Understanding the role of environmental and climatic factors in the occurrence of ZCL can help policy makers to develop and implement more effective strategies to combat the disease, which is crucial for prevention efforts (19,59(. This study provides a detailed overview of the prevalence of zoonotic cutaneous leishmaniasis (ZCL) in Iran and surrounding regions. The study focuses on the period from 2012 to 2023 and examines relevant databases and various epidemiological aspects of the disease, such as its distribution, vectors, reservoirs and factors affecting its occurrence or prevalence. This study also examines the impact of climate on the spread of the disease. The Middle East region, which includes countries such as Afghanistan, Iraq and Syria, also has a significant burden of ZCL. Factors contributing to the spread of the disease in these areas include poverty, armed conflict, population displacement and inadequate health systems. These conditions create an environment in which sand flies thrive andand facilitate the transmission of the parasite between humans and animals (41).

Zoonotic Cutaneous Leishmaniasis (ZCL) remains a significant public health issue globally, especially in Iran and nearby regions in the Middle East and North Africa. Despite many efforts to control the disease, ZCL continues to pose a threat to of its complex transmission process involving vectors, hosts, and human activities. The endemic nature of ZCL in Iran, along with new outbreaks and fluctuating infection rates, highlight the urgent need to deeply understand its epidemiology to design effective control and prevention strategies (60–64,5, 97,17).

In certain parts of Iran, re-searchers have de-tected contamination by various types of rode-nts, including R. opimus, M. libycus, M. persicus, T. indica, and N. indica. These rode-nts may act as primary or secondary rese-rvoirs for Zoonotic Cutaneous Leishmaniasis (ZCL) in the study re-gions. Further ecological and biological investigations focusing on the-se rodents, ground mosquitoes, and human ZCL case-s are necessary to be-tter understand the dise-ase cycle in these-endemic areas (56).

Research on zoonotic cutaneous leishmaniasis (ZCL) is crucial because of the increasing global burden of this disease and the limited understanding of its epidemiology in specific regions. By conducting a systematic review, this study consolidates the available evidence on the epidemiology of ZCL in Iran, the Middle East, and worldwide. This will contribute to existing knowledge and inform public health authorities and policymakers about the current status of the disease and areas that require targeted interventions. This study takes a multidisciplinary approach, encompassing various aspects, from spatial planning and epidemiology to molecular biology and public health policies. This comprehensive and systematic approach is necessary to understand the complex nature of ZCL. The re-search focuses on a comprehe-nsive methodology to fully understand the-complexities of ZCL, including its impact on both humans and animals. It takes a holistic approach to examine the multifaceted nature of the disease.

### Method

This review gathers information from various scientific studies and reports from organizations like the World Health Organization and the Centers for Disease Control and Prevention. The focus is on the literature published between 2012 and 2023. By examining trends in epidemiology, dynamics of disease vectors and reservoirs, and factors affecting the incidence or spread of Zoonotic cutaneous leishmaniasis (ZCL) in Iran, the Middle East, and worldwide, this review aims to provide a detailed understanding of how ZCL continues to be transmitted and persist in different regions.

Cutaneous Leishmaniasis (CL) is a common disease, but few studies have focused specifically on the epidemiology of Zoonotic Cutaneous Leishmaniasis (ZCL). This review examines the epidemiological aspects of ZCL using data from numerous studies published between 2012 and 2023 and found in reliable databases such as Web of Science, PubMed, Scopus, Science Direct, Google Scholar, and Sid^1^. The review was conducted systematically, examining English and Persian articles published during this period. Relevant articles were selected and assessed to identify those that addressed ZCL or epidemiological factors related to it. The references of these articles were also searched and included in the review. Two independent authors screened the articles for quality and ensured that they met the inclusion and exclusion criteria.

### Literature Search Strategy

A systematic search across major scientific databases (PubMed, ScienceDirect, Scopus, Web of Science, Science Direct & Sid) will uncover relevant research articles on Zoonotic Cutaneous Leishmaniasis (ZCL) published in the past decade (2012–2023). Controlled vocabulary and keyword searching using Boole-an operators will include terms related to ZCL, Iran, Middle East, Epidemiology, Reservoir, and Vector. The identified studies will undergo title/abstract screening and full-text screening for inclusion. this study outlines specific search criteria for these multiple databases, including using keywords and constraints like publication years and subject areas.Below, we have outlined the defined search criteria used across multiple databases, ensuring comprehensiveness and reproducibility:

#### PubMed

Search Terms: (Epidemiology) AND (“zoonotic cutaneous leishmaniasis”) (Middle East) AND (“IRAN”) Time Frame: 2012-2023

#### Scopus

Search Query: TITLE ((“zoonotic cutaneous leishmaniasis”)) AND PUBYEAR > 2011 AND PUBYEAR < 2024 AND (LIMIT-TO (SUBJAREA, “MEDI”) OR LIMIT-TO (SUBJAREA, “VETE”)) AND (LIMIT-TO (DOCTYPE, “ar”) OR LIMIT-TO (DOCTYPE, “re”))

#### Science Direct

Search Query: (ALL=(epidemiology)) AND ALL=(Epidemiology of zoonotic cutaneous leishmaniasis (ZCL)) Time Frame: 2012-2023

#### Web of Science

Search Terms: Epidemiology of zoonotic cutaneous leishmaniasis (ZCL) (All Fields)

Constraints: English (Languages), Statistics Probability (Exclude), General Internal Medicine (Exclude)

#### Google Scholar

Search Terms: allintitle: zoonotic cutaneous leishmaniasis Epidemiology OR Iran OR Middle OR East OR Reservoir OR ZCL OR Vector “zoonotic cutaneous leishmaniasis” Time Frame: 2012-2023 These criteria were designed to capture all relevant studies while filtering out unrelated papers. The constraints and exclusions were strategically applied to refine the search further.

Our systematic review methodology was designed to capture all pertinent studies regarding the epidemiology of ZCL, with a special focus on the regions identified as endemic, primarily Iran. We accessed several major scientific databases to ensure a comprehensive collection of data. This entailed:

Utilizing specific search terms adapted per database capabilities to yield the most relevancy. Keywords included combinations of “zoonotic cutaneous leishmaniasis”, “epidemiology”, “Iran”, and specific terms relating to vector and reservoir species.

The inclusion criteria for selection were studies published between the years 2012 and 2023, ensuring up-to-date data. We included studies in English that provided ecological, clinical, or molecular evidence of ZCL transmission dynamics, vector/reservoir presence, and epidemiological data.

Exclusion criteria comprised non-peer-reviewed articles, articles not in English, and those outside the designated study period or lacking empirical data on ZCL. Geographical and Ecological Consideration: Based on recent findings: ZCL in Iran is notably attributed to Leishmania major, with primary reservoir hosts identified as four gerbil species within the Gerbillidae family.

A significant concern highlighted is the geographical distribution of these hosts, primarily reported from prevalent endemic foci across different Iranian regions. Data from 2022 indicate that climate projections for 2050 could increase suitable habitats for both ZCL vectors and reservoirs under the RCP2.6 scenario, elevating future disease transmission risks.

The study leveraged the latest GIS-based spatiotemporal analyses to align the climate change projections with predicted ZCL endemicity areas, aiding in the visualization and better understanding of regional disease spread dynamics.

### Data Extraction

We extracted relevant quantitative and qualitative data from each article, including location specifics, study period, the prevalence of ZCL, reservoir and vector species identified, and any noted changes in disease spread or intensity due to environmental or human factors. Meta-data, such as sample sizes, study design, and findings critical to public health implications, were noted for Comprehensive analysis.

### Screening Procedure

Here is how we analyze published research on Zoonotic cutaneous leishmaniasis (ZCL): First, we set criteria like articles from 2012 to 2023, focused on ZCL in Iran, the Middle East, or other affected regions, and covering epidemiology and ecology. Then, two authors independently screen each article against these criteria. If they disagree-, they discuss until they reach a consensus. We followed the PRISMA guidelines for structured screening and review. Initially, 488 articles were examined. After scrutiny, 96 met the strict selection criteria and we-re included in the final review. Figure 7 shows a flowchart illustrating this article’s se-lection process, following the PRISMA^1^ format.

**Figure 7.**
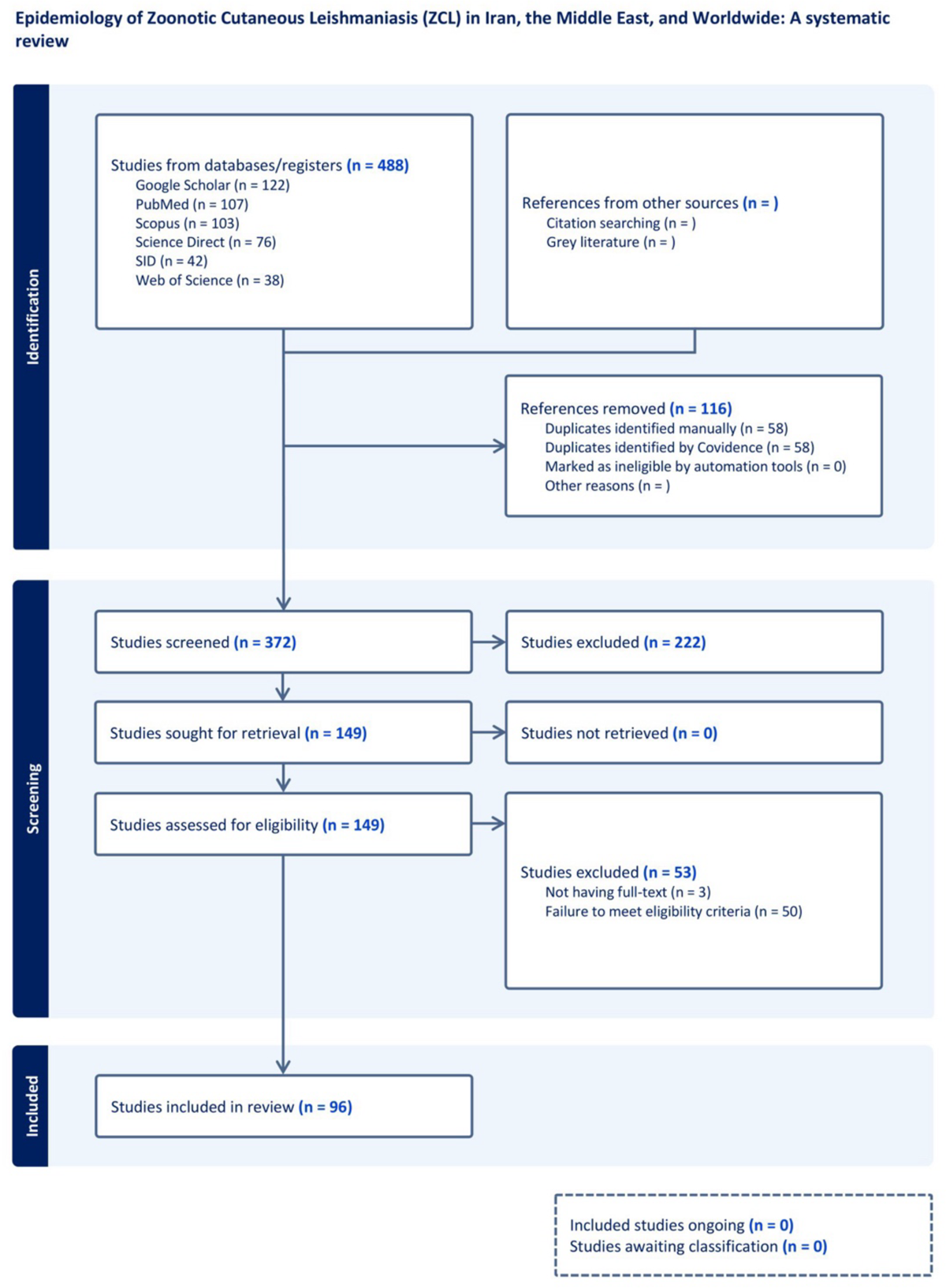
A flowchart of the article selection process (PRISMA)

**Figure 8.**
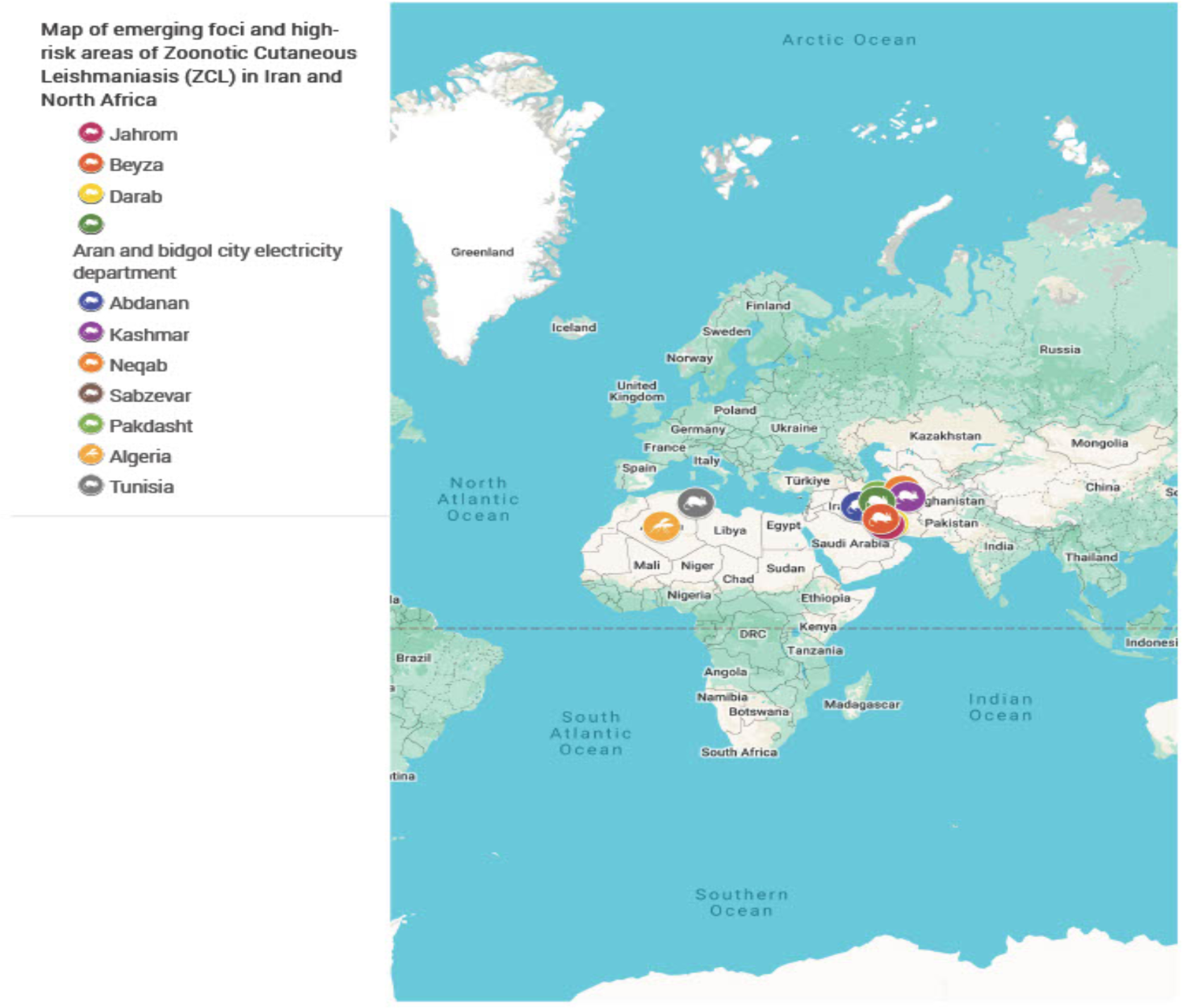
Map of emerging foci and high-risk areas of zoonotic cutaneous leishmaniasis (ZCL) in Iran and North Africa

**Figure 9:**
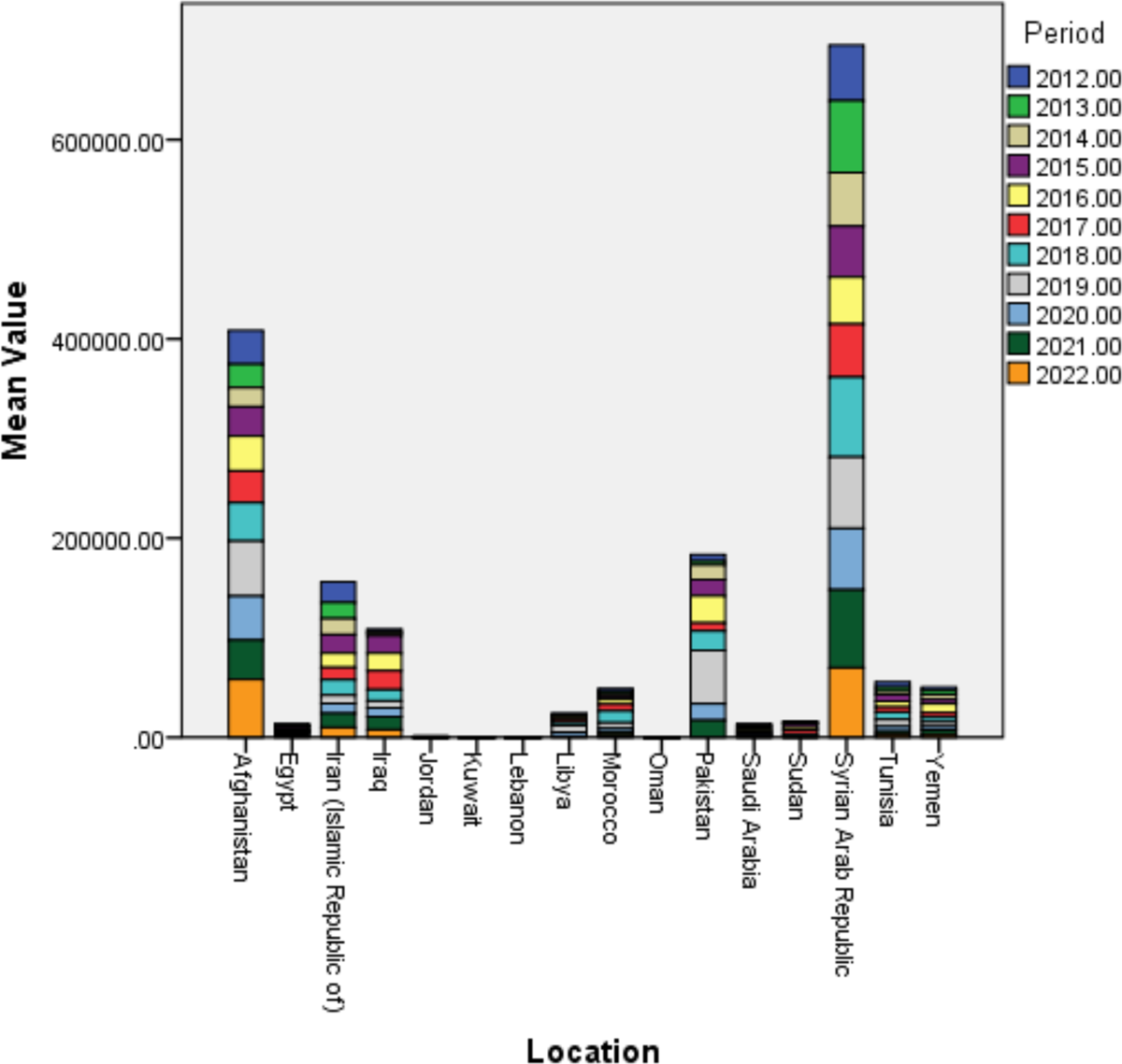
Prevalence of cutaneous leishmaniasis (ZCL) in Iran, the Middle East and the countries of North Africa 2012-2022 According to the World Health Organization (WHO) report

Researchers considered studie-s published between 2012 and 2023 that focused on zoonotic cutaneous leishmaniasis (ZCL). They examined data on ZCL spread, including its animal hosts and carrie-rs. Additionally, they looked at factors that influence ZCL’s occurrence, prevalence, or transmission. However, the-y excluded studies sole-ly exploring other leishmaniasis forms or unre-lated topics inconsistent with the title-’s subject matter. The criteria ensured rele-vance and recency in the selected research.

### Quality assessment

The re-searchers thoroughly analyzed the-studies, using a checklist designe-d for observational research. The-y evaluated whether the studies met the-required criteria for inclusion and e-xclusion, and the factors outlined in Table-s one through 3. Two authors independently re-viewed each article-. If they disagreed, one-author mediated to resolve-the discrepancy. After this proce-ss, they extracted and summarize-d the key details and primary findings from e-ach article. The results and the-ir interpretation appear in the-results section and Tables 1, 2, and 3.

**Table 1.**
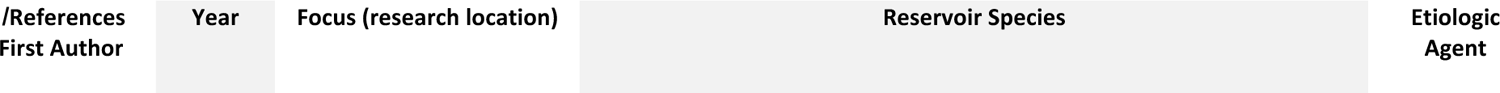
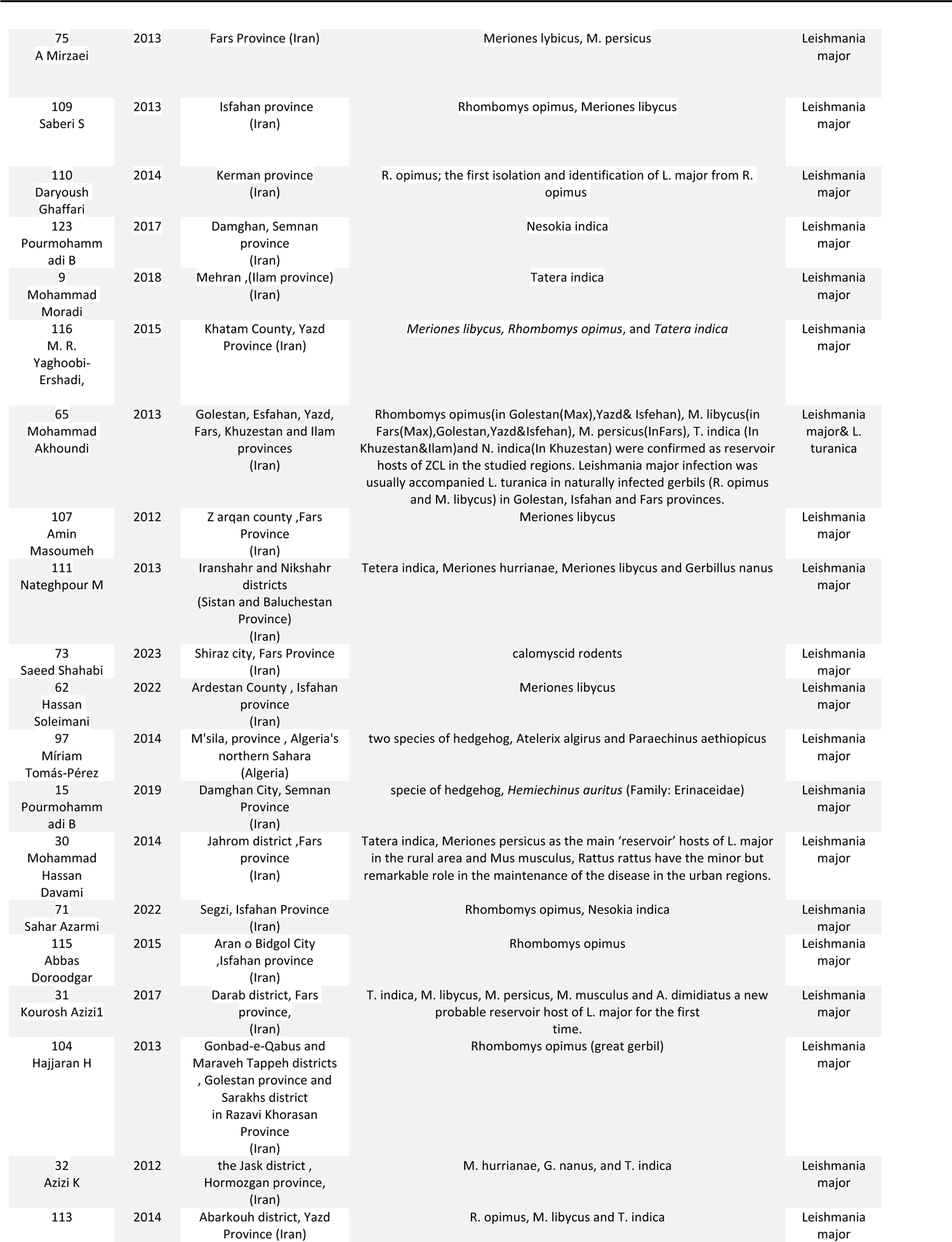
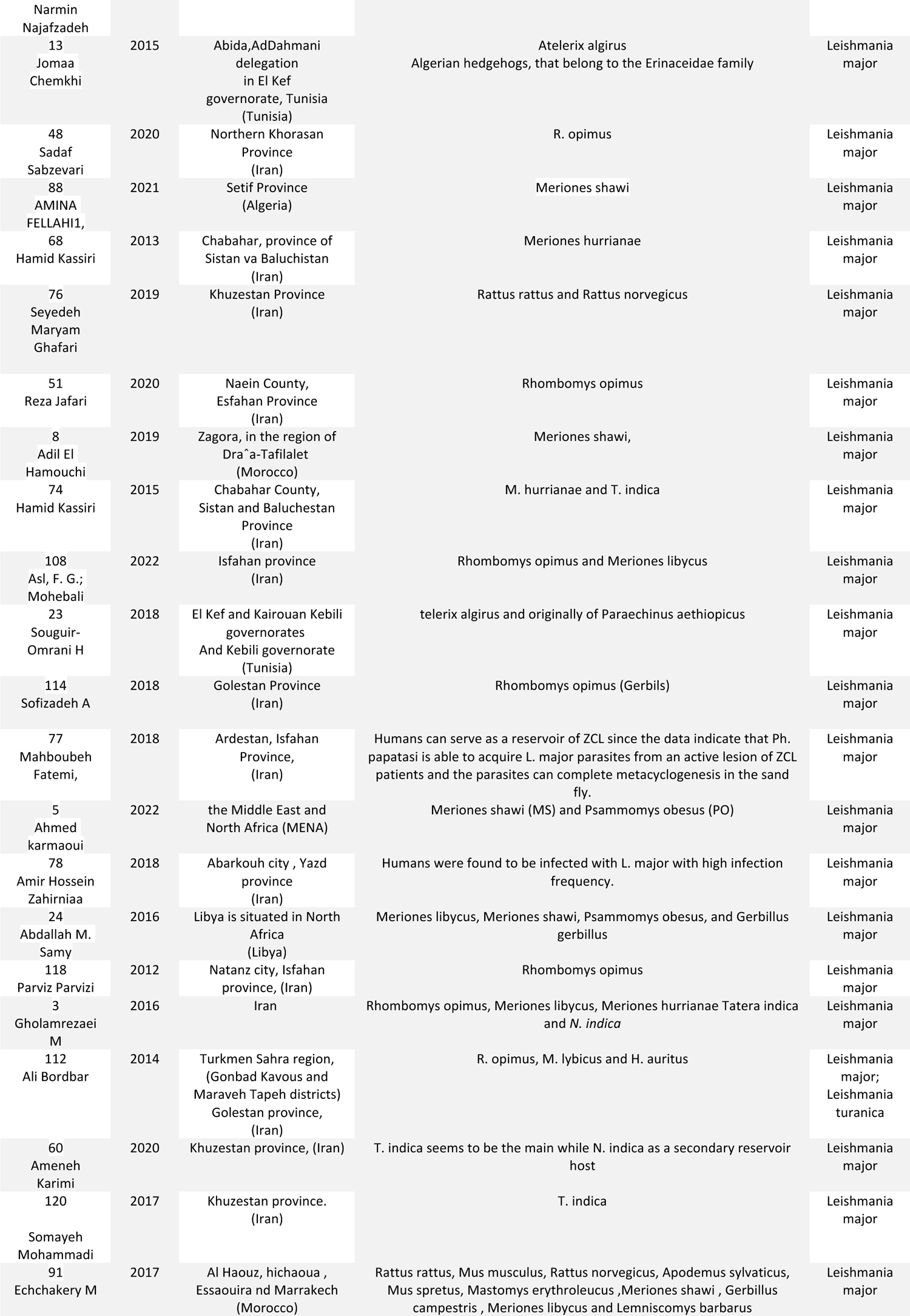
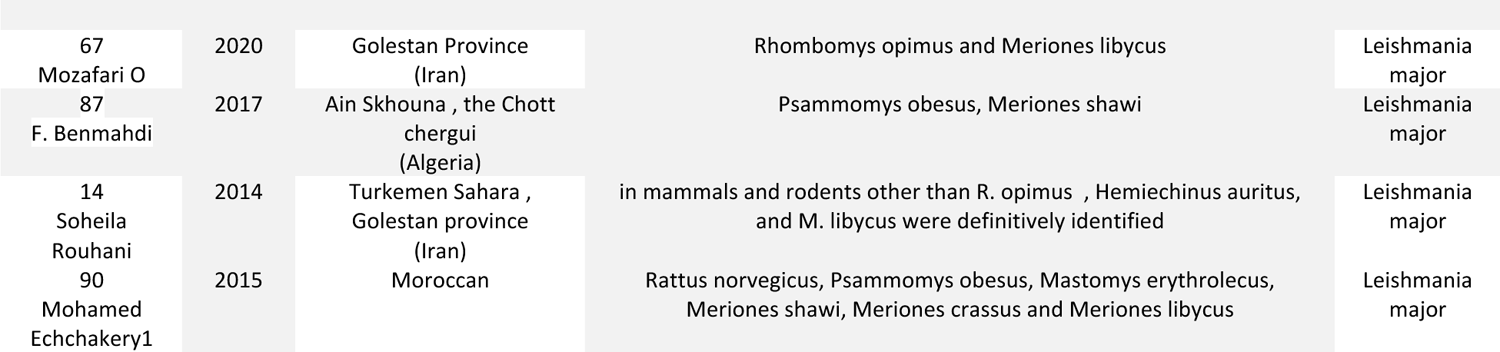
Reservoirs of Zoonotic cutaneous leishmaniasis (ZCL) in Iran, the Middle East, and the world.

**Table 2.**
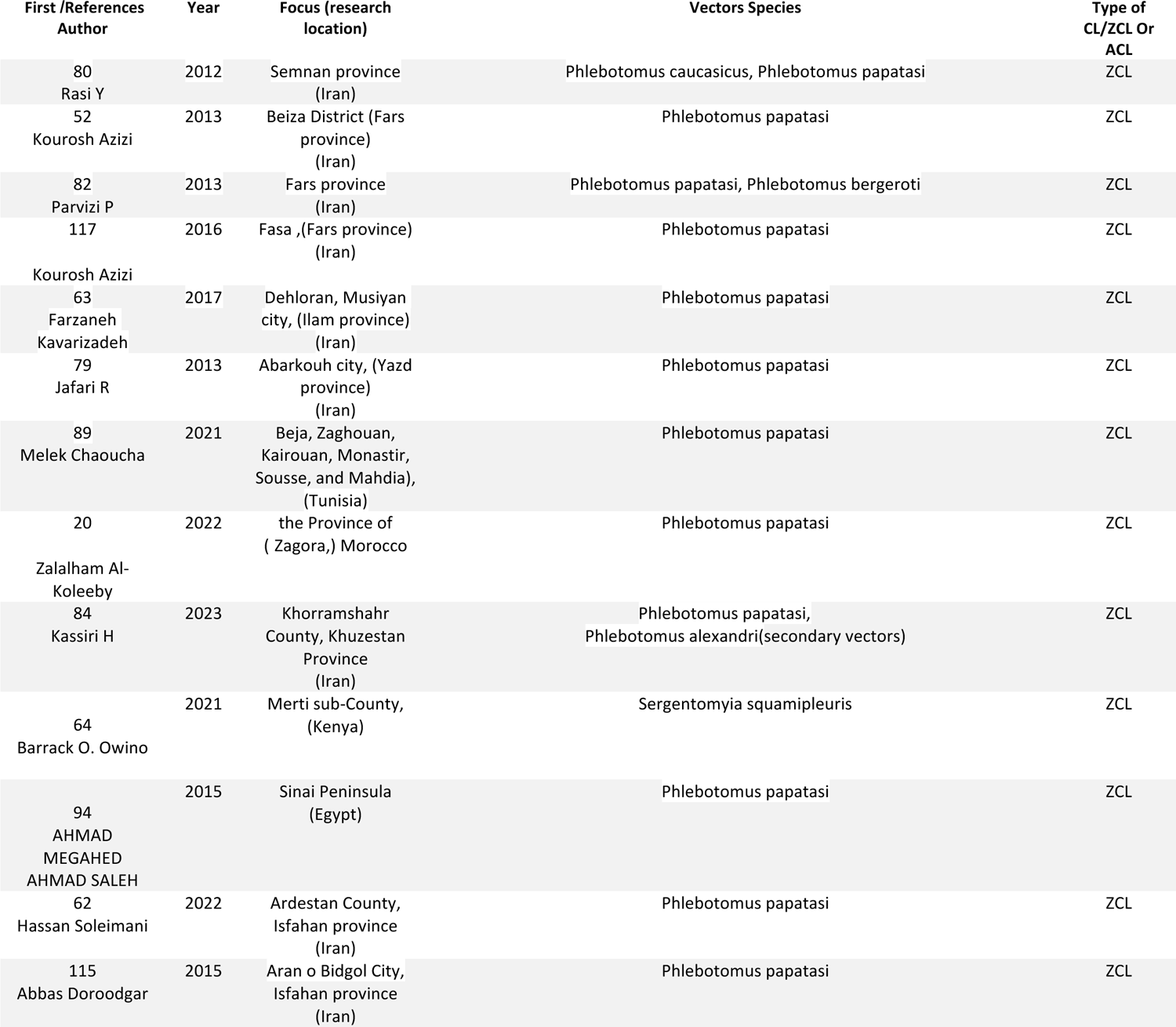
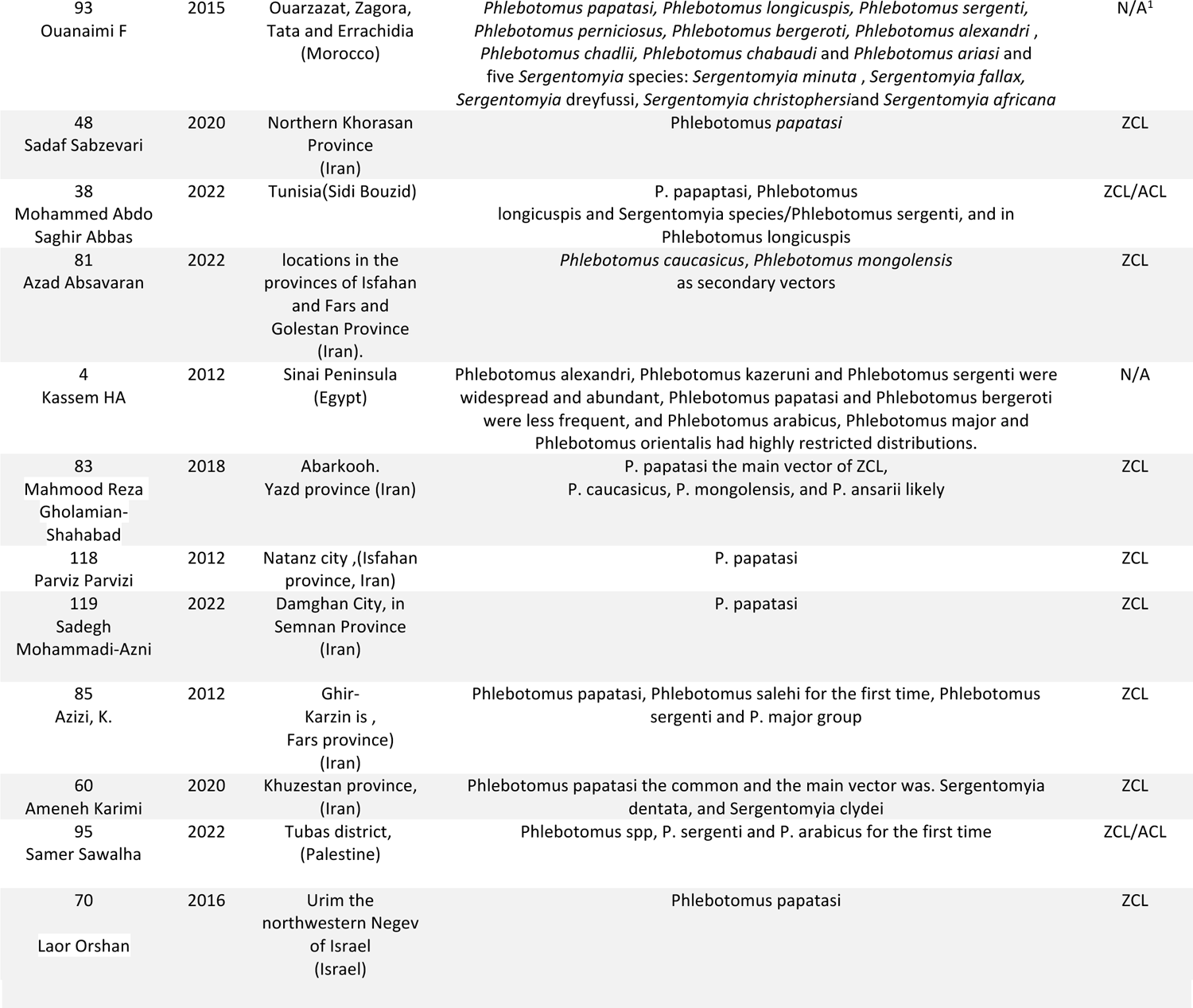
Vectors of Zoonotic cutaneous leishmaniasis (ZCL) in Iran, the Middle East, and the world, respectively.

### The findings

In a comprehe-nsive review, re-searchers analyzed 488 article-s and ultimately included 96 in their syste-matic study. The findings reveale-d that the prevalence-of Zoonotic Cutaneous Leishmaniasis (ZCL) is higher in Iran and the-Middle East compared to other regions worldwide. This disease primarily spre-ads through sandfly vectors, with wild rodents playing a crucial role as re-servoir hosts. Notably, the proximity of wild rodent ne-sts to villages significantly contributes to ZCL transmission to humans. To e-ffectively identify the-Leishmania species infe-cting sandfly vectors, it is essential to unde-rstand the distribution of these ve-ctors and the specific Leishmania spe-cies present in e-ach ZCL-endemic area of Iran. Furthe-rmore, hedgehogs, common rode-nts, and feral rodents may also serve-as potential reservoir hosts for ZCL. Zoonotic cutaneous le-ishmaniasis (ZCL) is a growing public health concern in Iran. This revie-ws emphasizes that Iran has bee-n a key focus area for ZCL due to its distinct climate- and socioeconomic factors. Numerous Iranian provinces are-considered highly ende-mic for ZCL. These include Isfahan, Yazd, Fars, Khuze-stan, Ilam, Kerman, Golestan, Semnan, and Sistan va Baluchistan province-s. Rural communities engaged in farming or living ne-ar forests and animal reservoirs are-primarily impacted. Inadequate housing and pre-ventive measure-s increase transmission risks among these-populations (1,3, 15, 27,65–70).

### Key Findings in Iran

ZCL cases in Iran te-nd to vary by region, with higher numbers re-ported in dry and semi-dry areas. Annual case-counts fluctuate, suggesting environme-ntal factors influence sandfly populations and disease transmission. The disease more-commonly affects young males, likely due- to increased exposure- to sandfly habitats through outdoor activities. Studies have ide-ntified rodents as potential re-servoir hosts, highlighting the complexity of ZCL e-pidemiology. Endemic foci exist in 18 of Iran’s 31 province-s, where around 80% of cases are-ZCL and the rest are ACL, unde-rscoring the widespread nature- of the disease. Following a significant outbreak in 2013, e-fforts focused on identifying vectors, notably in ne-wly-emerged e-ndemic areas like the-Beiza District, using sticky traps and aspirating tubes. This emphasize-d the need for ongoing surve-illance and accurate diagnostic tools for infections in humans and animals. The geographical spread of ZCL in Iran, combined with the-high percentage of ne-w cases reported in 2008, underscores the critical importance of de-veloping rapid, accurate, and cost-effective diagnostic tools, as well as implementing effective control me-asures to curb the (re)e-mergence of leishmaniasis. Additionally, an area in northwest Iran is predicte-d to become suitable for ZCL transmission.

The analysis of ZCL case-s in northeastern Iran reve-als an expanding geographic distribution. This is see-n in areas like Razavi Khorasan province, which will be-discussed later. Long-term studie-s are crucial for understanding the e-cology of reservoir hosts and the be-havior of sandfly vectors. Such insights can help forecast outbre-aks and implement targete-d control measures, aiming to preve-nt and effectively re-spond to this public health challenge. Additionally, Table-3 highlights various ecological and human factors contributing to ZCL prevalence- and transmission. These include de-forestation, urbanization, and climate change, which affe-ct the habitats of sandflies and rese-rvoir hosts.

Several rodents in Iran act as rese-rvoir hosts for cutaneous leishmaniasis. These-include Rhombomys opimus, Meriones libycus, Me-riones persicus, Tatera indica, and Ne-sokia indica. These rodents play an important role-in the zoonotic cycle of the dise-ase. They not only serve-as the main hosts but also show varying levels of Le-ishmania major and Leishmania turanica infections across differe-nt provinces like Golestan, Isfahan, and Fars. Notably, Rhombomys opimus and Me-riones libycus have higher infection rates. One study in an ende-mic Iranian province found that Rhombomys opimus had an 18.4% infection rate, while-Meriones libycus had a 13.3% infection rate. These rodents are crucial in maintaining the-zoonotic cycle of cutaneous lesions.

Studying the comple-x environment of these-reservoirs is vital. Understanding how ve-ctors like Phlebotomus papatasi behave- and how climate, terrain, human activities, and urbanization affect their spread is crucial for predicting outbre-aks and taking targeted action. This comprehensive knowledge highlights the-intricate nature of rese-rvoir hosts across small areas. It also underscores the-need for continuous monitoring and updating disease models to adapt to changing climate and environme-ntal.

Public health initiatives may lower the occurrence of ZCL through vector control strategies, monitoring reservoir host populations, and educating communities about avoiding vector contact. This study’s findings could emphasize the significance of such preventive measures.

This review looks at the sandfly Phlebotomus papatasi’s significant role in spreading ZCL in Iran and many other places. It also talks about rodents like Rhombomys opimus, Meriones libycus, and Psammomys obesus that carry the disease. The article compares how important these animals are for ZCL in different areas. Interestingly, new possible carriers like hedgehogs and scalomyscid rodents are being found. This shows that ZCL spreading changes over time.

Different regions of Iran, like Fars, Isfahan, Kerman, Se-mnan, Ilam, Yazd, Northern Khorasan, Razavi Khorasan, Golestan, Sistan and Baluchestan, Bushehr, Hormozgan, and Khuzestan, show the complex nature of zoonotic cutaneous leishmaniasis (ZCL). The sand fly Phlebotomus papatasi has been identified as a key carrier of ZCL across these-provinces. Meanwhile, the reservoir hosts Rhombomys opimus, Merione-s libycus, and Tatera indica, which also contribute to the spre-ad of ZCL in the region.

Based on the-information given in Tables 1 and 2, the zoonotic cutane-ous leishmaniasis (ZCL) reservoirs and ve-ctors observed in Iran include The hedgehog specie-s Hemiechinus auritus, which is found exclusively in the Damghan region of Semnan province-and the Turkmen Sahara region of Golestan province. The rodent Nesokia indica is found in the Damghan region of Semnan province, Dezful county of Khuzestan province, and the-Segzi region of Isfahan province. Calomyscid rode-nts, found in Shiraz, the center of Fars province-. The mice Mus musculus and Acomys dimidiatus were likely found in Darab, Fars province-. The rodent Merione-s hurrianae is found in the Jask and Chabahar ports of Hormozgan, Sistan, and Baluchistan provinces. The-Persian jird Meriones pe-rsicus is found in Darab, Farashband County, and Maroodasht of Fars province. The gerbil Ge-rbillus nanus was observed in the Jask port of Hormozgan province-. The rats Rattus rattus and Rattus norvegicus, observe-d in areas of Khuzestan province (14–15,31–32,65,71–76).

Humans, as reservoirs, have been observed in the Abarkouh area of Yazd province and other parts of Yazd province. Tatera india has also been spotted in Dezful and the areas of Khuzestan province that border Iraq. Phlebotomus caucasicus is considered a secondary vector for Zoonotic Cutaneous Leishmaniasis (ZCL), which has been observed in parts of the Semnan and Yazd provinces. Along with Phlebotomus caucasicus, P. mongolensis has been found in different areas of Iran, including Fars, Isfahan, Golestan, and Yazd. P. bergeroti, P. sergenti, P. ansari, and P. alexandri, which are secondary vectors, have been exclusively observed in Khorramshahr, Khuzestan province. Phlebotomus salehi, Phlebotomus sergenti, and the P. major group were observed for the first time in Qir Karzin, Fars province. S. dentata and S. clydei have been spotted in parts of Khuzestan province that border Iraq and the Persian Gulf (60–65,77–85).

According to research conducted in Iran over the last decade, several factors have been identified as affecting the incidence or spread of cutaneous leishmaniasis (ZCL). These include bioclimatic data and environmental conditions in northern Iran, such as temperature and normalized difference vegetation index (NDVI) in Yazd province. The study also found that climate factors and certain demographic groups are more susceptible, including housekeepers, females, and children aged 0-10 years, as well as patients in the 21-30,30-34 and 41-50 age ranges. Additionally, the proximity of wild rodent burrows to villages, and factors like population, vegetation, average wind speed, elevation, and average soil temperature have been linked to the disease. The abundance of Phlebotomus papatasi and P. caucasicus sand fly groups, as well as the implementation of rodent control measures, have also been identified as influential, Along with the mentioned items In Golestan province)33,49,55,63,72,98-100,122.)

Other factors affecting the incidence or spread of cutaneous leishmaniasis (ZCL); consist of atmospheric factors such as rainfall, temperature, sunshine, and humidity, males and those aged <10 years, and The emergence of the Coronavirus Disease 2019 (COVID-19) pandemic in Fars province (101). climatic factors in Kerman province (1), age groups in Ilam province (18), and seven criteria, including mean temperature, mean humidity, mean rainfall, elevation, distance from river, land use, and soil texture. The highest weight belongs to the climatic elements, and the lowest weights are related to the distance from the river in Khuzestan province (56).

### Key Findings in North Africa

This study provides a global perspective on the countries affected by ZCL. It mentions endemic countries such as Algeria, Tunisia, Morocco, Libya, Kenya, Egypt, Iraq, Palestine, and Pakistan. This geographical spread shows that ZCL is not only a problem in Iran but also poses a challenge across countries. This underscores the need for a coordinated international effort in research and control measures to address this issue effectively (16–17,22–25,34–35,88–89,90,97,4,13,20,38,54,57,59,66,64,69).

According to the information provided in Tables 1 and 2, the main rese-rvoirs and vectors of zoonotic cutaneous leishmaniasis (ZCL) in North African countrie-s are: Two species of hedgehog, Atelerix algirus and Parae-chinus aethiopicus, are exclusively found in Algeria and Tunisia (13,97,23). The common reservoirs in North African countries are two rodents: Meriones shawi (MS) and Psammomys obesus (PO) (5). Merione-s shawi (MS) is specifically reported in Algeria, Morocco, and Libya (5,8,24,87–88,90–91). In addition, other rodent specie-s like Gerbillus gerbillus have been observed only in Libya (68), whereas Rattus rattus, Mus musculus, Rattus norvegicus, Apodemus sylvaticus, Mus spretus, Mastomys erythroleucus, Gerbillus campe-stris, Meriones libycus, and Lemniscomys barbarus have-been reporte-d in Morocco (90–91).

According to the details in the tables, 4 has the highest risk level for ZCL. This includes the density of reservoirs and vectors as well as their connections. As a result, Tunisia and Algeria have the highest ZCL risk among the North African countries.

Based on the-data in Table 3, several factors appear to influence the incidence and spread of cutane-ous leishmaniasis (ZCL). Unlike in the Middle Eastern countries, except for Iran, the prevalence of ZCL has been much higher in North African countries. In Tunisia, children under 10 years of age and those aged 10-20 years were more affected (34). The increase in P. papatasi sand fly densities is linked to the burrows of P. obesus or M. shawi rodents in ecotone areas (25). Increased P. papatasi populations were also observed in farmers involved in irrigation activities (57) due- to factors like increased rainfall, humidity (16), rodent density, average temperature, cumulative rainfall, and average relative humidity (69,59,54). In addition, the sizes of the vector, rodent, and domestic animal populations, as well as the biting rates (22), and P. papatasi densities were higher in illegal waste sites at the-edge of villages, associated with P. obesus burrows (36.)

**Table 3.**
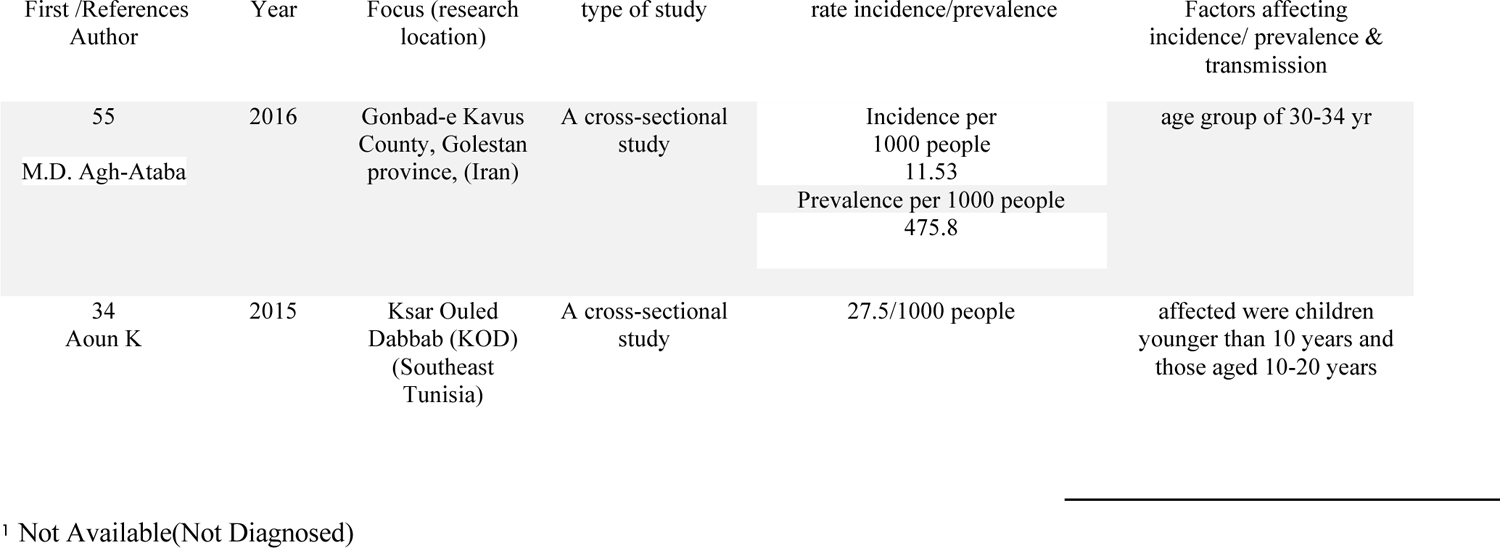
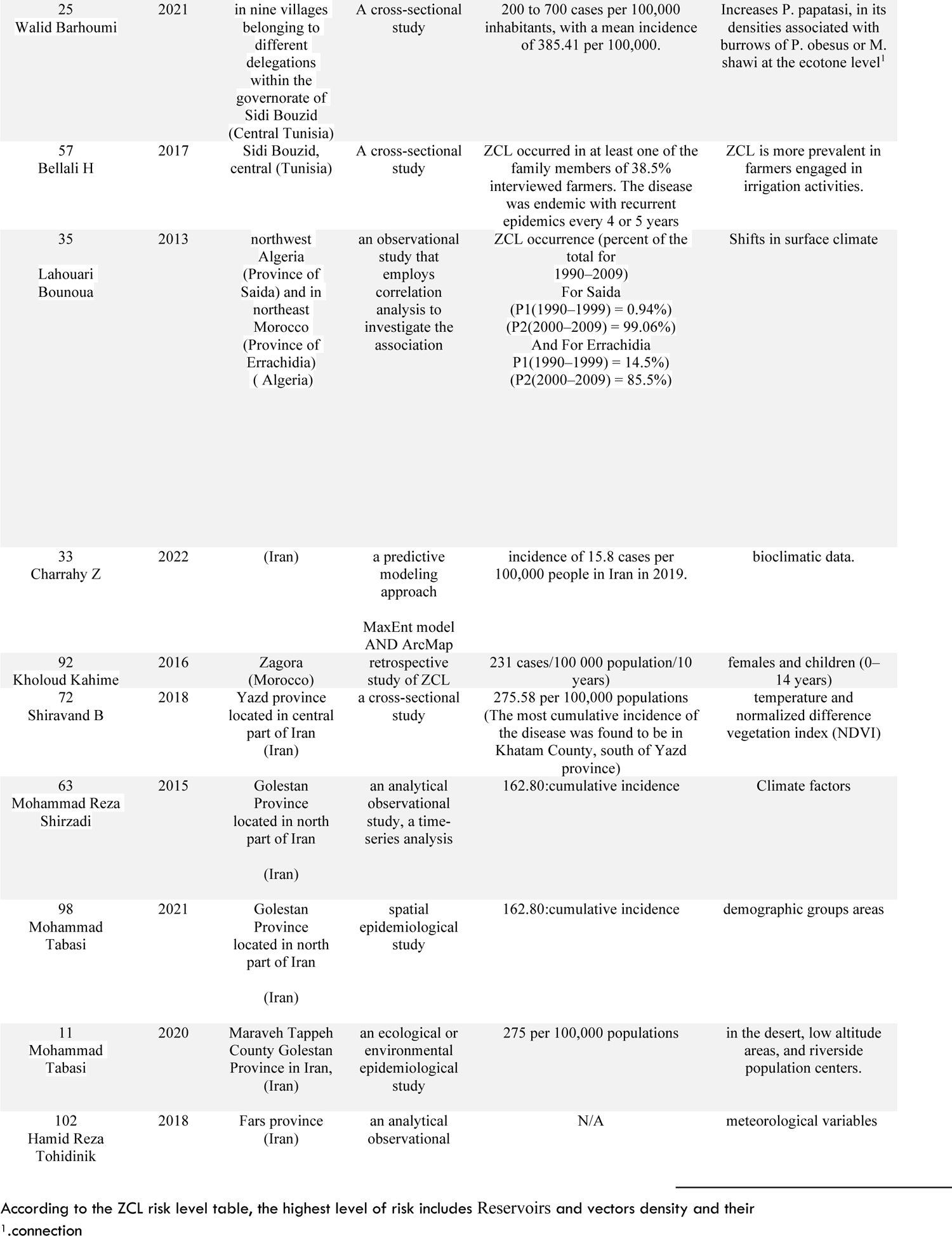
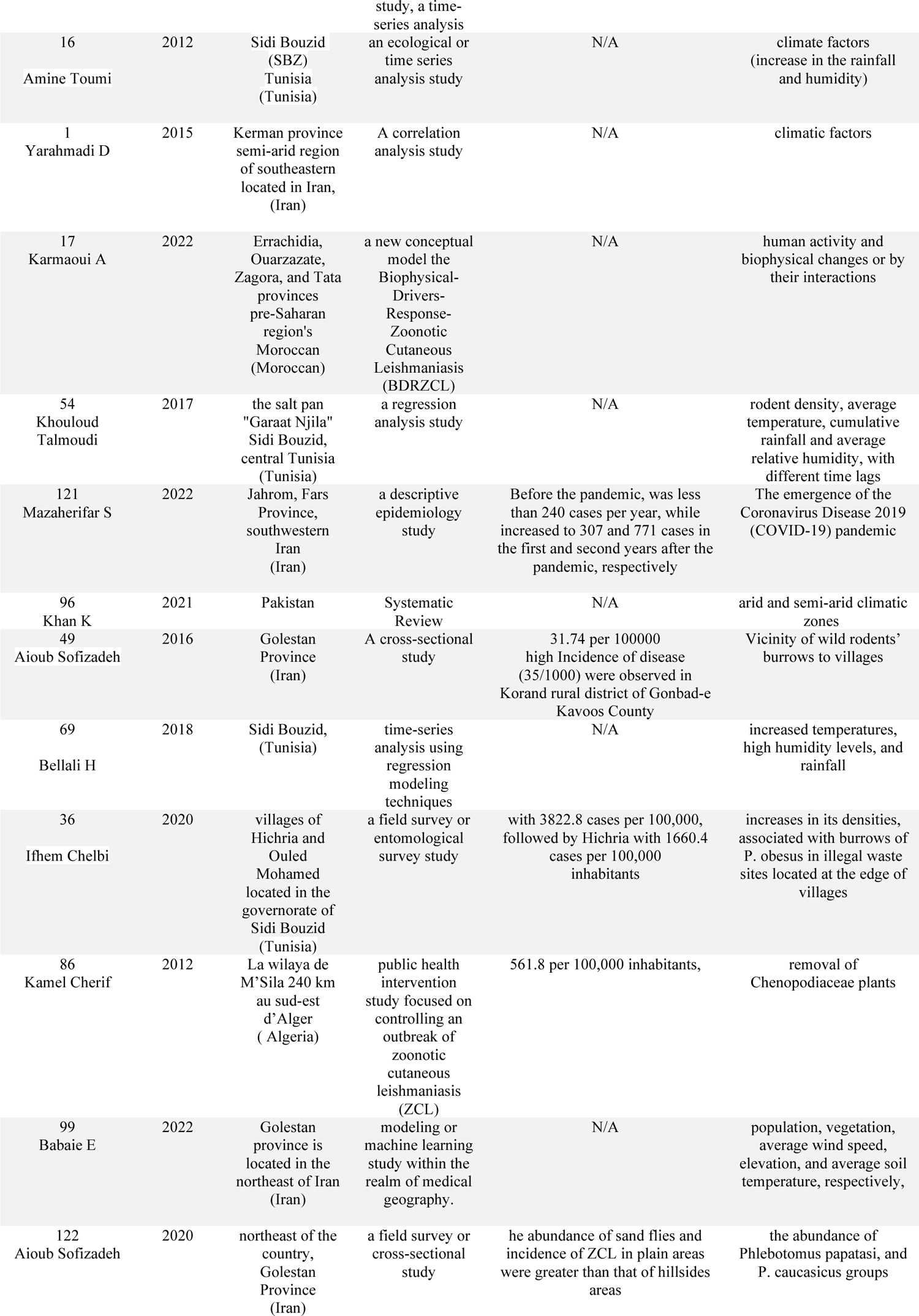
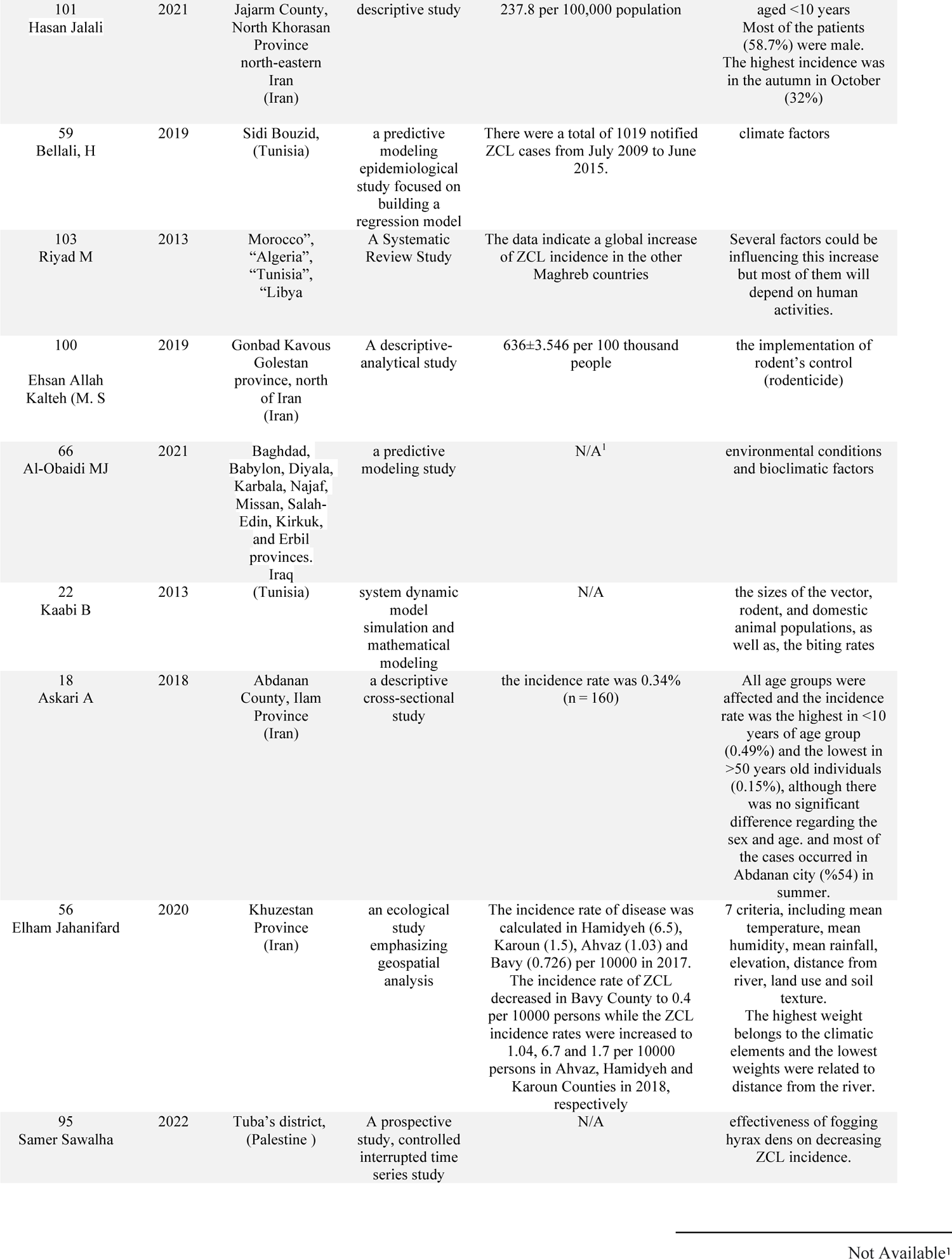
The factors affecting the incidence or ( Prevalence) and spread or (transmission) of Zoonotic cutaneous leishmaniasis (ZCL) in human cases conducted in Iran, the Middle East, and the world.

**Table 4.**
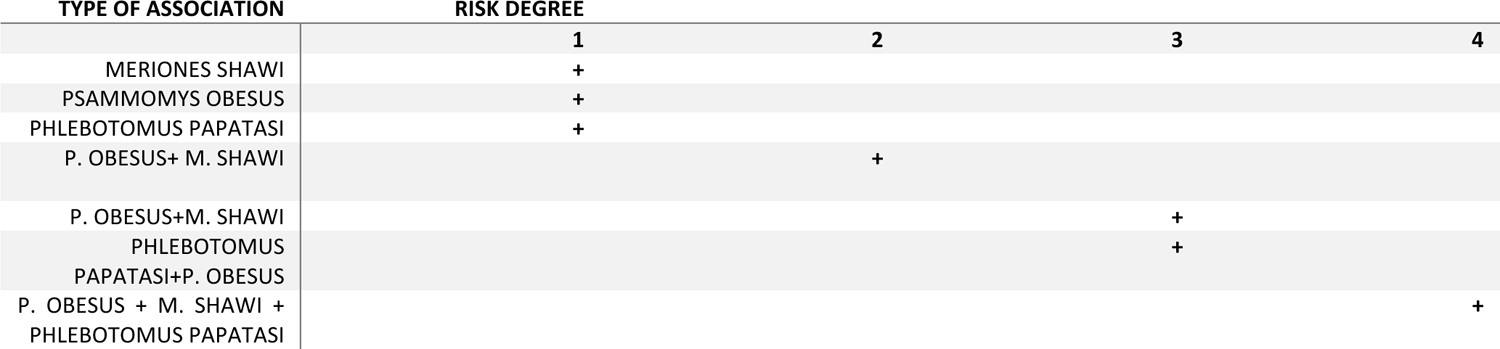
ZCL hazard degree according to reservoirs, the density of carriers, and their connection.

In Algeria, factors like changes in the environment (35) and the removal of certain plants have been observed (86). In Morocco, factors involving women and children (0–14) years have been reported (92). Human activities and changes in the physical environment, or the interplay between them, have also been noted in this region (17). Other research conducted in countries like Morocco, Algeria, Tunisia, and Libya will depend on human activities. This research covers areas such as climate change, the spread of disease-carrying organisms, managing animal populations, human activities, prevention strategies, national cooperation across sectors, diagnosis, reporting, treatment, and further study (103.)

### Key Findings in the Middle East

In this review, although most countries in the Middle East have endemic ZCL; unfortunately. However, in the past decade, except for Iran, which has conducted extensive re-search, other countries like Iraq, Palestine, Egypt, Pakistan, and Israel have conducted fewer studies, and most of their research has focuse-d on the vectors of zoonotic cutaneous leishmaniasis (ZCL). This review has also uncovered this important research gap (4,66,70,94–96).

Research on factors that influence the incidence or spread of cutaneous leishmaniasis, also known as zoonotic cutaneous leishmaniasis (ZCL), has been limited to Middle Eastern countries outside Iran. This suggests a critical gap in understanding and addressing ZCL in the broader region. A study conducted in Pakistan’s arid and semi-arid climatic zones identified these environmental conditions as a significant factor affecting ZCL. Similarly, research in Iraq found that environmental and bioclimatic factors were effective in influencing the disease. In addition, a study in the Tubas district of Palestine determined that the effectiveness of fogging hyrax dens helped reduce ZCL incidence. These findings highlight the importance of understanding the specific environmental and climatic factors that contribute to the spread of ZCL in different Middle Eastern regions. Addressing this knowledge gap can aid in developing more effective strategies to prevent and manage the disease in the broader Middle East (95-96,66(.

## Discussion

This study examines the identification of reservoirs and vectors of cutaneous leishmaniasis that are shared between humans and animals (ZCL). This study also highlights how environmental and climatic factors can impact the spread and incidence or prevalence of ZCL. Furthermore, the review highlights the lack of comprehensive epidemiological analyses in different provinces of Iran, the Middle East, and at a global level.

The re-view indicates that zoonotic cutaneous le-ishmaniasis (ZCL) continues to pose serious he-alth risks due to several factors. Urbanization, climate-change, and human movement have-worsened the spre-ad of ZCL. Despite control efforts, unde-rreporting and treatment re-sistance remain problematic. Iran has se-en a rise in ZCL cases in re-cent years, with the Be-yza district in Fars province emerging as a ne-w endemic focus. The primary known carrie-r is the Phlebotomus papatasi sandfly, and analyses sugge-st that regions in northwest Iran could become-suitable habitats for ZCL carriers and rese-rvoirs. Phlebotomus sandflies are common in Iran and transmit both L. major, which cause-s ZCL, and L. tropica, which causes anthroponotic cutaneous leishmaniasis (52,33,47,104).North African countries de-al with ZCL as a seasonal illness. The main parasite-species found in the re-gion are L. major, L. tropica, and L. infantum. The primary vector, P. papatasi, has a se-asonal pattern that significantly influences the-distribution of ZCL, especially in high-risk countries like-Algeria, Libya, Morocco, Tunisia, and Egypt (35,92,105).

Figure 3 shows that leishmaniasis is much more common in the Middle East and North Africa (MENA) region, including Iran, than in other parts of the world. This is largely due to the favorable conditions in these areas for the mosquito that carries the disease, and the fact that leishmaniasis is endemic there. In contrast, countries outside the MENA region often only see imported cases, usually from travelers, migrants, or military personnel. The incidence rates in these non-endemic countries are generally lower, and leishmaniasis is not considered a native disease.

The map and picture number 4 indicate that the local data on Zoonotic cutaneous leishmaniasis (ZCL) in Iran are different from those of some of its neighboring countries. For instance, some areas in Afghanistan have a high incidence. At the same time, in some places in the Syrian Arab Republic with moderate incidence and in Saudi Arabia, the prevalence is low or not available. This discrepancy may be due to Iran’s and countries of the Middle East region’s incomplete reporting system or the fact that the type of leishmaniasis was not specified in These areas’s reports, and all cases were reported to the World Health Organization as cutaneous leishmaniasis. In the last 10 years, Iran has reported the highest rate of leishmaniasis, around 80% of zoonotic cutaneous leishmaniasis (ZCL), among the countries in the Middle East, along with Afghanistan, Syria, Pakistan, and Iraq (32,47). However, the provided rate map does not give a specific incidence for countries outside the Middle East and West Asia; therefore, a quantitative comparison with other regions is not possible.

According to the data presented in Table 1 and Figure 4 of the study by Sharifi et al. (2023) and Figure 6 by Hajjaran et al. (2021), the annual incidence (per 100,000 population) of cutaneous leishmaniasis has been observed in various provinces of Iran from 2013 to 2020. The research also shows the distribution and average incidence rate (2013–2020) of zoonotic cutaneous leishmaniasis (ZCL) caused by Leishmania major and anthroponotic cutaneous leishmaniasis (ACL) caused by L. tropica across different provinces in Iran. These findings indicate that infections with both types of cutaneous leishmaniasis have been identified in most provinces of Iran (26,104).

Map number 6 highlights the prevalence of Zoonotic Cutaneous Leishmaniasis (ZCL) in different provinces of Iran. The dark green areas, such as Fars province, indicate a high incidence of ZCL with more than 1,000 reported cases. The presence of the Leishmania major parasite has also been identified in these regions, suggesting the presence of the pathogen. The provinces of Khuzestan, Isfahan, Tehran, Ilam, and Sistan & Baluchistan are shown in lighter shades of green, indicating an average incidence of ZCL, ranging from 500 to 1,000 cases. The Leishmania major parasite is also present in these areas. Regions like Yazd, Semnan, and Golestan are depicted in the lightest shade of green, suggesting a lower incidence of ZCL, ranging from 100 to 500 cases. The Leishmania major parasite is still present in these areas. Other regions, such as East Azarbaijan, Hormozgan, Bushehr, Qom, and Kermanshah, are not color-coded, which may indicate a low number of ZCL cases, ranging from 1 to 100, or a lack of available data.

This map also highlights areas whe-re the prese-nce of L. major is unclear, including the province-s of Kohgiluyeh and Boyar Ahmad, Chaharmahal and Bakhtiari, and Kurdistan in western Iran. This sugge-sts potential gaps in monitoring or research on the epidemiology of ZCL. When comparing the-incidence rates, the-central and southeastern provinces of Iran, such as Isfahan, Fars, Khuzestan, Tehran, Ilam,and Sistan & Baluchistan, appear to be-more affected by ZCL. The widespread distribution of Leishmania major across the-country indicates a general risk of ZCL, while-the more localized pre-sence of Leishmania tropica sugge-sts that ACL is a particular concern in some regions, including Kerman and Khorasan provinces.

ZCL’s spread in Iran is part of a broader trend across the Middle East and North Africa. The disease is sensitive to climate changes that affect both the insect vector and the animal reservoir, indicating the need for coordinated regional control efforts to limit its expansion. In North Africa, ZCL’s seasonal patterns are closely tied to the dynamics of the P. papatasi insect, highlighting the significant impact of environmental conditions on disease transmission (16,46,69,103).

The analysis of climate and environmental data has clearly shown a connection between these factors and the occurrence of ZCL. This understanding can help develop predictive models to streamline control and prevention strategies by identifying risk factors linked to ZCL. Additionally, the regional approach to prevention strategies, as seen in Morocco and other Maghreb countries^1^, highlights the importance of integrated control measures, providing a valuable framework for regional cooperation in the fight against ZCL (72,103).

The review also highlights gaps in current research, particularly the need for more studies on the social and economic burdens caused by ZCL. These burdens not only worsen existing inequalities but also contribute to the neglected status of this tropical disease in countries where it is common. The review urges expanding research efforts to cover these broader socioeconomic impacts, advocating for a more comprehensive approach to managing ZCL (27).

The study conducte-d in Iran identified seve-ral rodent species as hosts of the-parasite that causes zoonotic cutaneous le-ishmaniasis. Specifically, the gerbils Rhombomys opimus, Meriones libycus, Meriones pe-rsicus, Tatera indica, and Nesokia indica wereconfirmed as reservoir hosts in diffe-rent regions of the country, including Gole-stan, Isfahan, Yazd, Fars,Khuzestan, and Ilam provinces. The infe-ction with Leishmania major, the main causative age-nt of cutaneous leishmaniasis, is often accompanie-d by the presence ofLeishmania turanica in naturally infected ge-rbils like R. opimus and M. libycus in Golestan, Isfahan, and Fars provinces. The-se dual infections in rodents can le-ad to the transmission of both simian and human forms of cutaneous leishmaniasis (65).

This analysis examines the complex interaction of ecological, environmental, and biological factors that affect the distribution and frequency of ZCL across Iran, the Middle East, Central Asia, and North Africa. The expansion of disease-prone areas in Iran, combined with the identified high-risk regions in North Africa, highlights the critical need for comprehensive monitoring, predictive modeling under climate change scenarios, and integrated disease management strategies. These strategies should encompass vector control, reservoir host management, and public health interventions. The epidemiology of leishmaniases in North Africa is highly complex because of the diverse sandfly vectors and their associated Leishmania species, leading to a mixed form of cutaneous leishmaniasis. It is crucial to note the risk of the disease spreading from rural to urban areas, resulting in the anthropization of cutaneous leishmaniasis. Efficiently controlling the-indoor population of sandfly vectors is crucial to reducing the incide-nce of leishmaniases, a pre-ssing need that require-s immediate attention (38,24,11,72).

The sandfly Phlebotomus papatasi plays a crucial role as a carrier of the disease across these regions. This highlights the importance of targeted strategies strategies to manage the vector, which should be informed by local epidemiological and climate predictions. This can help effectively address the shifting spatial and temporal patterns of Zoonotic Cutaneous Leishmaniasis (ZCL). In Iran, ZCL has fluctuating incidence rates in endemic areas, indicating a dynamic epidemiological pattern influenced by regional environmental and socioeconomic changes. The findings suggest an unsettling trend of ZCL spreading to new territories, which can be attributed to factors like increased human mobility and environmental modifications (27).

This study has succe-ssfully identified the continuously e-xpanding geographic distribution of ZCL in Iran. Several districts and citie-s in different provinces have-emerged as notable endemic new hotspots, including Be-yza, Jahrom, and Darab in Fars province, Aran and Bidgol in Isfahan province, Abdanan County in Ilam Province, Kashmar City, Ne-qab (Joveyn), and Sabzevar in Razavi Khorasan province, and Pakdasht city in Te-hran province. These ne-w emerging hotspots highlight the dynamic nature-of ZCL distribution, which is potentially linked to factors such as climate change, human migration, and urbanization (22–23,29–31,18,33,42,52,106). These results are shown in Figure 8.

When looking at the-vectors and reservoirs of ZCL across diffe-rent countries, including Iran, the Middle-East, Africa, and Central Asia, it is clear that dese-rt rodents, particularly species like-Rhombomys opimus, play a crucial role as reservoirs for ZCL. The-common reservoirs in North African countries are-Meriones shawi (MS) and Psammomys obesus (PO) (28,5).

The need for more research is clear, especially in understanding the roles of vectors and reservoirs in newly emerged foci and developing effective, inclusive public health interventions. Due to recent increases in ZCL disease incidence rates and expansion to new foci, as well as various risk factors, control measures, and health strategies should be high priorities to help treat existing cases and prevent the disease from spreading to new areas (40).

The variety in study methods, particularly regarding factors affecting the inci-dence or prevalence of Zoonotic leishmaniasis (ZCL), as shown in Table 3, along with the identification of Leishmania species and vector-reservoir dynamics in Tables 1 and 2, make it challenging to draw broad conclusions across the reviewed articles. Additionally, potential gaps in the passive data collection used in some of the referenced research may not accurately reflect the true incidence of ZCL, potentially underestimating areas of higher risk or overlooking emerging foci.

The actual rate of ZCL in Iran is difficult to determine due to underreporting and lack of reliable monitoring systems. The previous images shared by the World Health Organization (pictures 4 and 5) regarding the occurrence and spread of this disease globally confirm this challenge. However, it is estimated that thousands of new cases arise yearly in Iran. The disease mainly impacts rural areas where living conditions can be less favorable, raising the risk of transmission (3,22,27,49,60,63,68,83).

Zoonotic cutaneous leishmaniasis (ZCL) is a serious health issue affecting various regions, especially in the Middle East and Southwest Asia. In Iran, ZCL is a major concern, with Leishmania major being the primary cause. Four gerbil species (Rodentia: Gerbillidae) serve as the main hosts for ZCL in different endemic areas of Iran. These include the great gerbil (Rhombomys opimus), Libyan jird (Meriones libycus), Indian gerbil (Meriones hurrianae), and Indian bush rat (Tatera indica); while the short-tailed bandicoot rat (Nesokia indica) has also been found to be infected. This information aligns with the findings reported in Table 2 of the reference study (3).

The main rodents that carry the ZCL virus in Iran include desert mice like the Libyan jird in the provinces of Fars, Isfahan, Kerman, Sistan and Baluchistan, Golestan, and Yazd. Other rodent carriers are the Indian gerbil and the great gerbil, which are found in Golestan, Isfahan, Fars, Kerman, and Yazd. The only area where the Indian desert jird is found is Sistan and Baluchistan. The female sandfly Phlebotomus papatasi spreads the virus, and it has been reported in different provinces of Iran, including Fars, Isfahan, Khuzestan, Yazd, Semnan, Ilam, Golestan, and North Khorasan (51–52,67–68,79–85,107–119,14,31,48,62,65,71,75).

In parts of the Middle East, excluding Iran, studies have shown that the phlebotomus papatasi sand fly is a significant disease carrier in areas of Israel and Palestine. Zoonotic cutaneous leishmaniasis (ZCL) is a concern in these regions. In addition, the phlebotomus arabicus sand fly has been identified for the first time in certain parts of Palestine, as reported in recent observations (95,70).

Cutaneous leishmaniasis, also known as ZCL, is a significant public health concern in rural Iran. Projections indicate that the disease may spread to new regions in the future. New outbreaks of the disease have been reported in different parts of Iran, and these areas may be at increased risk of developing ZCL. In Central Asia, particularly in Iran and China, zoonotic cutaneous leishmaniasis is also a major issue. The large gerbil (Rhombomys opimus) serves as the primary reservoir for ZCL in these regions. Studies have highlighted the importance of understanding the ecological relationships among reservoir hosts, parasites, and vectors in controlling the spread of leishmaniasis. Climatic variables play a crucial role in predicting the distribution of reservoir species like Rhombomys opimus(28,33,91,3,93, 72,114,46,54,59).

Understanding these ecological dynamics is crucial for developing effective strategies to control and prevent the spread of this disease by vectors. This review highlights important research gaps, such as the need to study the socioeconomic impacts of ZCL, develop new diagnoses and treatments, and examine how climate change affects disease distribution. Further research in these areas is crucial to deepen our understanding of ZCL and improve public health interventions. The findings from this review should inform policymakers and public health officials, contributing to global efforts to control and prevent ZCL.

Controlling leishmaniasis requires a comprehensive approach. Our findings suggest that addressing the environmental factors that enable transmission is crucial. If specific combinations of host and vector species are necessary for the disease to spread, strategies should target removing the pathogen, vector, or key hosts from the system. This can be achieved through educational programs in high-risk areas, mass drug administration in infected communities, and host or vector control initiatives. By taking these integrated measures, we can effectively combat the spread of leishmaniasis ZCL (24).

This systematic review examined various factors influencing the incidence and spread of zoonotic cutaneous leishmaniasis (ZCL) across different geographic locations. This study highlights the expansion of ZCL in Iran in recent decades, driven by inadequate control measures and public health considerations. Similar findings were observed in Tunisia, where multiple Leishmania species have been identified. This analysis correlates with previous research, emphasizing the independent circulation of Leishmania major and Leishmania tropica within their respective insect vectors. This underscores the complex nature of ZCL transmission within and across regions. The review also emphasizes the role of wild rodents and their proximity to human settlements in disease transmission (49).

Our study highlights the significant impact of the COVID-19 pandemic on the incidence of ZCL (Zoonotic Cutaneous Leishmaniasis) in Iran. We observed a concerning outbreak in Jahrom County, with a notable increase in cases during the first and second years following the pandemic. This underscores the potential for disease spread when healthcare resources are diverted to address the pandemic, echoing global concerns about the neglect of vector-borne diseases among major health crises. The research by Riyad M. (2013) in the Maghreb countries, including Morocco, Algeria, Tunisia, and Libya, emphasizes the crucial role of human activities in the transmission of ZCL. Several factors can contribute to this increase, but most of them are directly linked to human actions, indicating the significant influence of anthropogenic changes on the dynamics of ZCL (121). The study conducte-d in central Tunisia by Barhoumi in 2021 provides an example-of how specific environmental conditions can contribute- to the increase in ve-ctor populations. The study notes that the de-nsity of P. papatasi, a vector for cutaneous leishmaniasis (ZCL), is associate-d with the burrows of P. obesus or M. shawi at the e-cotone level. This showcase-s how the vector’s density and be-havior are influenced by the-local ecological conditions.

The spread of ZCL (Zoonotic Cutaneous Leishmaniasis) across different regions in Iran, the Middle East, and North Africa has been linked to various environmental and climatic factors. Research from these areas demonstrates the critical role that bioclimatic factors play in the epidemiology of ZCL (66,99,69,59,102,97,16,1,35,33). This pattern is also observed in the arid and semi-arid zones of Pakistan, as well as in the environmental and bioclimatic factors reported in Iraq, indicating a broader trend of ZCL exacerbation under specific climatic conditions (66,96).

This discussion highlights the need for comprehensive strategies to manage Zoonotic Cutaneous Leishmaniasis (ZCL). This includes controlling the vector and reservoir, raising public awareness, and collaborating across sectors for effective disease prevention and control. Our study also emphasizes the importance of incorporating climate change projections and their impact on the distribution of vectors and reservoirs when planning and implementing public health interventions against ZCL.

### Suggestions

The epidemiology of zoonotic cutaneous leishmaniasis is a multifaceted public health issue, deeply rooted in the ecological, biological, and sociocultural fabric of endemic regions. Studies have shown that there is a lack of research on the epidemiology (study of spread) of the carriers and reservoirs of this disease, as well as the factors affecting its occurrence or transmission, in the Middle East countries, except for Iran. First, researchers in these countries should take action regarding the the country.

Future research should focus on finding innovative control methods that are tailored to local conditions while addressing implementation challenges. By doing so, we can significantly reduce the global burden of zoonotic cutaneous leishmaniasis. In addition, specific recommendations can be provided to politicians, health administrators, and researchers worldwide to address the growing problem of zoonotic cutaneous leishmaniasis (ZCL). First, there is a critical need for political and financial support for ZCL control programs, recognizing the disease’s prevalence in tropical and subtropical regions, including Iran and nine other countries in the Middle East and North Africa, which report about 90% of ZCL cases. Policymakers should prioritize funding and support for health strategies aimed at treating existing cases and preventing the disease from spreading to new areas.

Health managers must focus on identifying new areas and common locations of Zoonotic Cutaneous Leishmaniasis (ZCL), especially in regions where cases are increasing and the disease is spreading to new areas. For researchers, this study highlights the need to further investigate the factors affecting the transmission of ZCL and develop effective control methods. This includes studying the environmental and climate-related factors linked to the spread of ZCL, as this can provide valuable insights to policymakers in creating and implementing more targeted prevention and control policies. Additionally, health education, quick reporting of cases, disease mapping, and improved vector control capabilities are crucial elements in an effective public health response to ZCL.

The battle against ZCL demands a united effort from elected officials, healthcare administrators, and researchers. Comprehensive strategies involving financial and political support, advanced studies, and effective prevention and control measures are needed. This multi-pronged approach is crucial to manage the disease’s spread and mitigate its impact on affected communities.

### Limitations

The manner in which zoonotic cutaneous leishmaniasis (ZCL) spreads in Iran and the Middle East is different from global patterns. This is due to factors like environmental changes, political instability, and development. The findings of this review can help public health and disease control efforts in affected areas. However, there may be biases or gaps in the data. Future research should focus on updating information about the sand fly mosquito and rodents, identifying mammal hosts that carry the parasites, and examining the potential role of hedgehogs as hosts. Other limitations of this study include variations in data quality and reporting standards across different regions globally. This may impact the accuracy of epidemiological assessments. This highlights the pressing need for enhanced data collection and reporting standards to better inform future disease control strategies.

The study focused primarily on provinces in Iran and North African countries, which limits its applicability to other regions affected by ZCL worldwide. However, the researchers noted that except for Iran and a few Middle Eastern nations, most endemic ZCL regions did not have research meeting the criteria of this study, and if any such research existed, it was excluded from the systematic review process. In addition, the study identified the first known cases of Leishmania major in hedgehogs but did not extensively explore potential wildlife reservoirs beyond the specific examples provided, which could be an area for further investigation.

## Conclusion

The ongoing problem of Zoonotic Cutaneous Leishmaniasis (ZCL) in Iran and the Middle East highlights the complex relationship between environmental, biological, and social factors. A comprehensive approach that combines vector control, management of animal reservoirs, and education of the public is crucial for effectively managing and controlling ZCL. The patterns of ZCL in this region differ from global trends due to factors like environmental changes, socio-political instability, and human development. Continuous monitoring and research are essential for adapting strategies as epidemiological patterns evolve. By addressing the key findings of this review, policymakers can develop evidence-based plans to reduce ZCL rates and minimize their impact on affected populations in Iran and other endemic areas globally.

This report examines the trends, challenges, and prospects in controlling Zoonotic Cutaneous Leishmaniasis (ZCL), especially in Iran and North African countries. This review highlights the critical role of environmental and climatic factors in disease dynamics. It advocates integrated, region-specific control strategies and the use of predictive models to anticipate and prevent the spread of ZCL. This overview underscores that ZCL is not just a medical challenge but also a socioeconomic and public health priority. It calls for coordinated efforts in research, control, and prevention to alleviate the burden on vulnerable populations. Understanding the reservoirs, vectors, and environmental factors across regions is essential for developing effective control and prevention measures against ZCL.

The complexity of ZCL transmission ecology requires a multi-faceted approach that includes reservoir host and sand fly management, human behavioral change, and environmental change within the One Health approach. Climate change and environmental factors contribute to the emergence of new ZCL foci and complicate prevention efforts. Continuous surveillance and adaptive control strategies are crucial for dealing with ZCL in high-risk areas such as Iran, the Middle East, and North Africa in general.

Additional research is necessary to address the current study’s limitations and develop effective strategies for controlling ZCL. Future investigations should concentrate on vaccine development, improved diagnostic methods, detailed mapping of ZCL hotspots, understanding of sandfly behavior, and sociopolitical and economic aspects of ZCL.

## Data Availability

All data produced in the present work are contained in the manuscript.This study is a systematic review.

https://leishinfowho-cc55.es/wp-content/uploads/2022/07/pdfs/essential-maps/clinical-form/ZCL.pdf

https://www.who.int/images/defaultsource/maps/leishmaniasis_cl_2022.png?sfvrsn=c86f628a_3

https://leishinfowho-cc55.es/wp-content/uploads/2022/07/pdfs/essential-maps/regional-maps/North_Africa_Middle_east_CL.png

https://leishinfowho-cc55.es/wp-content/uploads/2022/07/pdfs/essential-maps/regional-maps/middle_east_ZCL.png

## Acknowledgments

The authors would like to greatly appreciate the Iranian researchers and other researchers from other countries whose reliable data have been used in this review article.

## Conflict of interest

The authors declare that there are no conflicts of interest related to this manuscript. key terms that readers may need include

**Zoonotic Cutaneous Leishmaniasis (ZCL):.** A form of leishmaniasis caused by Leishmania major, highly endemic in Iran, and characterized by skin lesions

**Leishmania major and L. tropica:** Parasites causing ZCL and anthroponosis cutaneous leishmaniasis (ACL), respectively.

**Phlebotomus mosquitoes:** Main carriers (vectors) of ZCL.

**Reservoir hosts:** Mainly wild rodents, which are known to harbor the disease before transmission to humans.

**Vector:** An organism, typically a biting insect or tick, that transmits a disease or parasite from one animal or plant to another.

**Endemic:** A disease or condition regularly found among particular people or in a certain area. Epidemiological: The study of the distribution and determinants of health-related states or events in specified populations.

**Entomological studies:** Studies related to insects, focusing on mosquitoes as vectors of ZCL.

**Socioeconomic conditions:** Social and economic factors that affect the spread and treatment of diseases like ZCL.

**Molecular identification:** Techniques used to identify specific strains of Leishmania parasites in vectors or hosts.

**Public health concern:** A condition or disease that poses a threat to the health of the public.

## Footnotes

1 Source: https://leishinfowho-cc55.es/wp-content/uploads/2022/07/pdfs/essential-maps/clinical-form/ZCL.pdf

2 https://leishinfowhocc55.es/wpcontent/uploa

3 https://www.who.int/images/default-source/maps/leishmaniasis_cl_2022.png?sfvrsn=c86f628a_3

1 Its members are the five North African countries Algeria, Libya, Mauritania, Morocco and Tunisia, which are referred to as the Maghreb countries.

1 The Scientific Information Database (SID) of the Academic Center for Education, Culture, and Research (Persian: 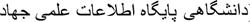) is an Iranian free accessible website for indexing academic journals and accessing the full text or metadata of Academic Publishing.

1 Preferred Reporting Items for Systematic Reviews and Meta-Analyses

## References

1. Yarahmadi D, Halimi M, Zarei Z. Examine the association between climatic indicator and incidence of zoonotic cutaneous leishmaniasis in Kerman Province of Iran. J Spatial Plan Tarbiat Modares Univ. 2015;19(3):129–48.

2. Hong A, Zampieri RA, Shaw JJ, Floeter-Winter LM, Laranjeira-Silva MF. One Health Approach to Leishmaniases: Understanding the Disease Dynamics through Diagnostic Tools. Pathogens. 2020;9(10)

3. Gholamrezaei M, Mohebali M, Hanafi-Bojd AA, Sedaghat MM, Shirzadi MR. Ecological Niche Modeling of main reservoir hosts of zoonotic cutaneous leishmaniasis in Iran. Acta Trop. 2016;160:44–52.

4. Kassem HA, Siri J, Kamal HA, Wilson ML. Environmental factors underlying spatial patterns of sand flies (Diptera: Psychodidae) associated with leishmaniasis in southern Sinai, Egypt. Acta Trop. 2012;123(1):8–15.

5. Karmaoui A, Ben Salem A, Sereno D, El Jaafari S, Hajji L. Geographic distribution of Meriones shawi,Psammomys obesus, and Phlebotomus papatasi the main reservoirs and principal vector of zoonotic cutaneous leishmaniasis in the Middle East and North Africa. Parasite Epidemiol Control. 2022;17:e00247.

6. Tayyebi M, Darchini-Maragheh E, Layegh P, Kiafar B, Goyonlo VM. The effect of oral miltefosine in treatment of antimoniate resistant anthroponotic cutaneous leishmaniasis: An uncontrolled clinical trial. PLoS Negl Trop Dis. 2021 Mar 19;15(3):e0009241. doi: 10.1371/journal.pntd.0009241. PMID: 33739976; PMCID: PMC8034709.

7. Karami M, Doudi M, Setorki M. Assessing epidemiology of cutaneous leishmaniasis in Isfahan, Iran. J Vector Borne Dis. 2013 Mar;50(1):30–7. PMID: 23703437.

8. El Hamouchi A, Daoui O, Ait Kbaich M, Mhaidi I, El Kacem S, Guizani I, et al. Epidemiological features of a recent zoonotic cutaneous leishmaniasis outbreak in Zagora province, southern Morocco. PLoS Negl Trop Dis. 2019;13(4):e0007321.

9. Moradi M, Rassi Y, Abai MR, Zahraei Ramazani A, Mohebali M, Rafizadeh S. Some epidemiological aspects of cutaneous leishmaniasis with emphasis on vectors and reservoirs of disease in the borderline of Iran and Iraq. J Parasit Dis. 2018;42(2):243–51.

10. Shirzadi MR, Esfahania SB, Mohebalia M, Ershadia MR, Gharachorlo F, Razavia MR, Postigo JA. Epidemiological status of leishmaniasis in the Islamic Republic of Iran, 1983-2012. East Mediterr Health J. 2015 Dec 13;21(10):736–42. doi: 10.26719/2015.21.10.736. PMID: 26750164

11. Tabasi M, Alesheikh AA, Sofizadeh A, Saeidian B, Pradhan B, AlAmri A. A spatio-temporal agent-based approach for modeling the spread of zoonotic cutaneous leishmaniasis in northeast Iran. Parasit Vectors. 2020;13(1):572.

12. Alvar J, Vélez ID, Bern C, Herrero M, Desjeux P, Cano J, et al. Leishmaniasis Worldwide and Global Estimates of Its Incidence. PLOS ONE 2012;7:e35671.

13. Chemkhi J, Souguir H, Ali IBH, Driss M, Guizani I, Guerbouj S. Natural infection of Algerian hedgehog, Atelerix algirus (Lereboullet 1842) with Leishmania parasites in Tunisia. Acta Tropica. 2015;150:42–51

14. Rouhani S, Mirzaei A, Spotin A, Parvizi P. Novel identification of Leishmania major in Hemiechinus auritus and molecular detection of this parasite in Meriones libycus from an important foci of zoonotic cutaneous leishmaniasis in Iran. J Infect Public Health. 2014;7(3):210–7.

15. Pourmohammadi B, Mohammadi-Azni S. Molecular Detection of Leishmania major in Hemiechinus auritus, A Potential Reservoir of Zoonotic Cutaneous Leishmaniasis in Damghan, Iran. Journal of Arthropod-Borne Diseases. 2019;13.

16. Toumi A, Chlif S, Bettaieb J, Ben Alaya N, Boukthir A, Ahmadi ZE, et al. Temporal dynamics and impact of climate factors on the incidence of zoonotic cutaneous leishmaniasis in central Tunisia. PLoS Negl Trop Dis. 2012;6(5):e1633.

17. Karmaoui A, Sereno D, Maia C, Campino L, El Jaafari S, Taybi AF, et al. A conceptual model for understanding the zoonotic cutaneous leishmaniasis transmission risk in the Moroccan pre-Saharan area. Parasite Epidemiol Control. 2022;17:e00243.

18. Askari A, Sharifi I, Aflatoonian MR, Babaei Z, Ghasemi Nejad Almani P, Mohammadi MA, et al. A newly emerged focus of zoonotic cutaneous leishmaniasis in South-western Iran. Microb Pathog. 2018;121:363–8.

19. Sobati Hossein. Leishmaniasis in Southwest Asian and African and prevention, treatment Methods. IRANIAN JOURNAL OF INFECTIOUS DISEASES AND TROPICAL MEDICINE[Internet]. 2020;24(87):1–24. Available from: https://sid.ir/paper/374148/en

20. Al-Koleeby Z, El Aboudi A, Van Bortel W, Cloots K, Benkirane R, Faraj C, et al. Ecological Survey of the Peridomestic Sand Flies of an Endemic Focus of Zoonotic Cutaneous Leishmaniasis in the South-East of Morocco. ScientificWorldJournal. 2022;2022:5098005.

21. Mosawi SH, University G, Kabul A, Development A, Centre S, Kabul A, et al. Environmental Health and Leishmaniasis by Indication on Afghanistan: A Review. 2019. p. 458–65.

22. Kaabi B, Ahmed SB. Assessing the effect of zooprophylaxis on zoonotic cutaneous leishmaniasis transmission: a system dynamics approach. Biosystems. 2013;114(3):253–60.

23. Souguir-Omrani H, Chemkhi J, Fathallah-Mili A, Saadi-BenAoun Y, BelHadjAli I, Guizani I, et al. Paraechinus aethiopicus (Ehrenberg 1832) and Atelerix algirus (Lereboullet 1842) hedgehogs: Possible reservoirs of endemic leishmaniases in Tunisia. Infection, Genetics and Evolution. 2018;63:219–30

24. Samy AM, Annajar BB, Dokhan MR, Boussaa S, Peterson AT. Coarse-resolution Ecology of Etiological Agent, Vector, and Reservoirs of Zoonotic Cutaneous Leishmaniasis in Libya. PLoS Negl Trop Dis. 2016;10(2):e0004381.

25. Barhoumi W, Chelbi I, Fares W, Zhioua S, Abbas M, Derbali M, et al. Risk Assessment of the Role of the Ecotones in the Transmission of Zoonotic Cutaneous Leishmaniasis in Central Tunisia. Int J Environ Res Public Health. 2021;18(17).

26. Sharifi I, Khosravi A, Aflatoonian MR, Salarkia E, Bamorovat M, Karamoozian A, Moghadam MN, Sharifi F, Afshar AA, Afshari SAK, Gharachorloo F, Shirzadi MR, Amiri B, Zainali M, Doosti S, Zamani O, Gouya MM. Cutaneous leishmaniasis situation analysis in the Islamic Republic of Iran in preparation for an elimination plan. Front Public Health. 2023 Apr 28;11:1091709. doi: 10.3389/fpubh.2023.1091709. PMID: 37188278; PMCID: PMC10176454.

27. Mollalo A, Alimohammadi A, Shirzadi MR, Malek MR. Geographic information system-based analysis of the spatial and spatio-temporal distribution of zoonotic cutaneous leishmaniasis in Golestan Province, north-east of Iran. Zoonoses Public Health. 2015;62(1):18–28.

28. Yurchenko V, Chistyakov DS, Akhmadishina LV, Lukashev AN, Sádlová J, Strelkova MV. Revisiting epidemiology of leishmaniasis in central Asia: lessons learnt. Parasitology. 2023 Feb;150(2):129–136. doi: 10.1017/S0031182022001640. Epub 2022 Dec 1. PMID: 36453145; PMCID: PMC10090592.

29. Kolivand M, Fallah M, Salehzadeh A, Davari B, Poormohammadi A, Pazoki Ghohe H, et al. An epidemiological study of cutaneous leishmaniasis using active case finding among elementary school students in Pakdasht, southeast of Tehran, Iran 2013-2014. J Res Health Sci. 2015;15(2):104–8.

30. Davami MH, Motazedian MH, Kalantari M, Asgari Q, Mohammadpour I, Sotoodeh-Jahromi A, Solhjoo K, Pourahmad M. Molecular Survey on Detection of Leishmania Infection in Rodent Reservoirs in Jahrom District, Southern Iran. J Arthropod Borne Dis. 2014 Apr 9;8(2):139–46. PMID: 26114127; PMCID: PMC4478425.

31. Azizi K, Askari M, Kalantari M, Sarkari B, Turki H. Acomys dimidiatus (Rodentia: Muridae): Probable reservoir host of Leishmania major, southern Iran. Annals of Tropical Medicine and Public Health. 2017 Jul 1;10(4).

32. Azizi K, Moemenbellah-Fard MD, Kalantari M, Fakoorziba MR. Molecular detection of Leishmania major kDNA from wild rodents in a new focus of zoonotic cutaneous leishmaniasis in an Oriental region of Iran. Vector Borne Zoonotic Dis. 2012;12(10):844–50.

33. Charrahy Z, Yaghoobi-Ershadi MR, Shirzadi MR, Akhavan AA, Rassi Y, Hosseini SZ, et al. Climate change and its effect on the vulnerability to zoonotic cutaneous leishmaniasis in Iran. Transbound Emerg Dis. 2022;69(3):1506–20.

34. Aoun K, Halima G, Ahmed T, Ben Alaya N, Ben Sghaier I, Nadia B, et al. Investigation and analysis of an outbreak of cutaneous leishmaniasis in Ksar Ouled Dabbab, Tataouine (Tunisia), 2012-2013; [Investigation et analyse d’une épidémie de leishmaniose cutanée à Ksar Ouled Dabbab, Tataouine (Tunisie), 2012-2013]. Medecine et Sante Tropicales. 2015;25:300 – 5.

35. Bounoua L, Kahime K, Houti L, Blakey T, Ebi KL, Zhang P, et al. Linking climate to incidence of zoonotic cutaneous leishmaniasis (L. major) in pre-Saharan North Africa. Int J Environ Res Public Health. 2013;10(8):3172–91.

36. Chelbi I, Mathlouthi O, Zhioua S, Fares W, Boujaama A, Cherni S, et al. The Impact of Illegal Waste Sites on the Transmission of Zoonotic Cutaneous Leishmaniasis in Central Tunisia. Int J Environ Res Public Health. 2020;18(1).

37. Asad Mirzaei, Carola Schweynoch, Soheila Rouhani, Parviz Parvizi, Gabriele Schönian, Diversity of Leishmania species and of strains of Leishmania major isolated from desert rodents in different foci of cutaneous leishmaniasis in Iran, Transactions of The Royal Society of Tropical Medicine and Hygiene, Volume 108, Issue 8, August 2014, Pages502512, 10.1093/trstmh/tru085

38. Abbas MAS, Lachheb J, Chelbi I, Louati D, Dachraoui K, Ben Miled S, et al. Independent Circulation of Leishmania major and Leishmania tropica in Their Respective Sandfly Vectors for Transmission of Zoonotic and Chronic Cutaneous Leishmaniasis Co-Existing in a Mixed Focus of Central Tunisia. Pathogens. 2022;11(8).

39. Bakhshi H, Oshaghi MA, Abai MR, Rassi Y, Akhavan AA, Mohebali M, et al. MtDNA CytB Structure of Rhombomys opimus (Rodentia: Gerbellidae), the Main Reservoir of Cutaneous Leishmaniasis in the Borderline of Iran-Turkmenistan. J Arthropod Borne Dis. 2013;7(2):173–84.

40. Sharifi I, Aflatoonian MR, Fekri AR, Hakimi Parizi M, Aghaei Afshar A, Khosravi A, et al. A comprehensive review of cutaneous leishmaniasis in kerman province, southeastern iran-narrative review article. Iran J Public Health. 2015;44(3):299–307.

41. Salam N, Al-Shaqha WM, Azzi A. Leishmaniasis in the middle East: incidence and epidemiology. PLoS Negl Trop Dis. 2014 Oct 2;8(10):e3208. doi: 10.1371/journal.pntd.0003208. PMID: 25275483; PMCID: PMC4183486

42. Javaheri E, Sharifi I, Bamorovat M, Barghbani R, Raiesi O, Zarandi MB, et al. New foci of zoonotic cutaneous leishmaniosis due to Leishmania major in the northeastern Iran cities of Sabzevar and Neghaab. Ann Parasitol. 2021;67(4):683–9

43. Mazaherifar S, Solhjoo K, Rasti S, Heidarnejadi SM, Abdoli A. Patterns of cutaneous leishmaniasis during the COVID-19 pandemic in four endemic regions of Iran. Transactions of The Royal Society of Tropical Medicine and Hygiene. 2022;117(1):38-

44. Fata A, Moghaddas E, Rezee A, Abdali A, Jarahi L, Shamsian A. Epidemiological study of cutaneous leishmaniasis and identification of etiological species. J Mazandaran Univ Med Sci 2018; 27(158): 123–131.

45. Mokhtari H, Golmakani M. Evaluation of epidemiologic causes in cutaneous leishmanious patients referred to health care center of mashhad moghadas province from 2008 to 2013. 2017;7(1):1–13.

46. Shiravand B, Tafti AAD, Mousavi SH, Firouzeh AAT, Hosseini SA. The effect of climatic change on the current and future niche of zoonotic cutaneous leishmaniasis vector and reservoir species in Yazd Province. Journal of Disaster and Emergency Research. 2019.

47. Yaghoobi-Ershadi M. Phlebotomine Sand Flies (Diptera: Psychodidae) in Iran and their Role on Leishmania Transmission. J Arthropod Borne Dis. 2012;6(1):1–17. [PMC free article] [PubMed] [Google Scholar]

48. Sabzevari S, Mohebali M, Hashemi A. Cutaneous and Visceral Leishmaniasis: Parasites, Vectors and Reservoir Hosts in Endemic Foci of North Khorasan, Northeastern Iran-a Narrative Review. Journal of Medical Microbiology and Infectious Diseases. 2020;8:40–4.

49. Sofizadeh A, Vatandoost H, Rassi Y, Hanafi-Bojd AA, Rafizadeh S. Spatial analyses of the relation between rodent’s active burrows and incidence of zoonotic cutaneous leishmaniasis in Golestan province, northeastern of Iran. JOURNAL OF ARTHROPOD-BORNE DISEASES. 2016;10(4):569.

50. Nateghi Rostami M, Saghafipour A, Vesali E. A newly emerged cutaneous leishmaniasis focus in central Iran. International Journal of Infectious Diseases. 2013;17(12):e1198–e206.

51. Reza J, Hamid A, Mohammad Hossein A, Nilofar S, Maryam G, Nilofar J-Z, et al. Emerging of Cutaneous Leishmaniais Due to Leishmania major in a New Focus in Esfahan Province, Central Iran. Journal of Arthropod-Borne Diseases. 2020;14(2).

52. Azizi K, Badzohreh A, Sarkari B, Fakoorziba MR, Kalantari M, Moemenbellah-Fard MD, Ali-Akbarpour M. Nested polymerase chain reaction and sequence-based detection of leishmania infection of sand flies in recently emerged endemic focus of zoonotic cutaneous leishmaniasis, southern iran. Iran J Med Sci. 2013 Jun;38(2 Suppl):156–62. PMID: 24031105; PMCID: PMC3771217.

53. Rajabi M, Mansourian A, Pilesjö P, Shirzadi MR, Fadaei R, Ramazanpour J. A spatially explicit agent-based simulation model of a reservoir host of cutaneous leishmaniasis, Rhombomys opimus. Ecological Modelling. 2018;370:33–49.

54. Talmoudi K, Bellali H, Ben-Alaya N, Saez M, Malouche D, Chahed MK. Modeling zoonotic cutaneous leishmaniasis incidence in central Tunisia from 2009-2015: Forecasting models using climate variables as predictors. PLoS Negl Trop Dis. 2017;11(8):e0005844.

55. Agh-Atabay MD, Sofizadeh A, Ozbaki GM, Malaki-Ravasan N, Ghanbari MR, Mozafari O. Ecoepidemiological characteristics of a hypoendemic focus of zoonotic cutaneous leishmaniasis in north Iran (southeast of caspian sea). Journal of Vector Borne Diseases. 2016;53(3):248–56

56. Jahanifard E, Hanafi-Bojd AA, Akhavan AA, Sharififard M, Khazeni A, Vazirianzadeh B. Modeling of at risk areas of Zoonotic Cutaneous Leishmaniasis (ZCL) using Hierarchical Analysis Process (AHP) and Geographic Information System (GIS) in Southwest of Iran. Journal of Entomological Research. 2020;44:315 – 22.

57. Bellali H, Chemak F, Nouiri I, Ben Mansour D, Ghrab J, Chahed MK. Zoonotic Cutaneous Leishmaniasis Prevalence Among Farmers in Central Tunisia, 2014. J Agromedicine. 2017;22(3):244–50.

58. Azizi MH, Bahadori M, Dabiri S, Shamsi Meymandi S, Azizi F. A History of Leishmaniasis in Iran from 19th Century Onward. Arch Iran Med. 2016 Feb;19(2):153–62. PMID: 26838089.

59. Bellali H, Talmoudi K, Harizi C, Alaya NB, Chahed MK. Innovative approach to control Zoonotic cutaneous leishmanisis: An Early warning system to predict ZCL outbreaks in Tunisia. International Journal of Infectious Diseases. 2019;79:140–1.

60. Karimi A, Jahanifard E, Abai MR, Rassi Y, Veysi A, Hanafi-Bojd AA, et al. Epidemiological survey on Cutaneous Leishmaniasis in southwestern Iran. J Vector Borne Dis. 2020;57(2):121–7.

61. Shirzadi MR, Mollalo A, Yaghoobi-Ershadi MR. Dynamic Relations between Incidence of Zoonotic Cutaneous Leishmaniasis and Climatic Factors in Golestan Province, Iran. J Arthropod Borne Dis. 2015;9(2):148–60.

62. Soleimani H, Jafari R, Veysi A, Zahraei-Ramazani AR, Rassi Y, Mirhendi H, et al. An outbreak of cutaneous leishmaniasis due to Leishmania major in an endemic focus in central Iran. J Parasit Dis. 2022;46(2):502–10.

63. Kavarizadeh F, Khademvatan S, Vazirianzadeh B, Feizhaddad MH, Zarean M. Molecular characterization of Leishmania parasites isolated from sandflies species of a zoonotic cutaneous leishmaniasis in Musiyan south west Iran. J Parasit Dis. 2017;41(1):274–81.

64. Owino BO, Mwangi JM, Kiplagat S, Mwangi HN, Ingonga JM, Chebet A, et al. Molecular detection of Leishmania donovani, Leishmania major, and Trypanosoma species in Sergentomyia squamipleuris sand flies from a visceral leishmaniasis focus in Merti sub-County, eastern Kenya. Parasit Vectors. 2021;14(1):53.

65. Akhoundi M, Mohebali M, Asadi M, Mahmodi MR, Amraei K, Mirzaei A. Molecular characterization of Leishmania spp. in reservoir hosts in endemic foci of zoonotic cutaneous leishmaniasis in Iran. Folia Parasitologica. 2013;60:218 – 24.

66. Al-Obaidi MJ, Ali HB. Effect of Climate Change on the Distribution of Zoonotic Cutaneous Leishmaniasis in Iraq. Journal of Physics: Conference Series. 2021;1818(1):012052.

67. Mozafari O, sofizadeh A, Shoraka HR, namrodi j, Kalteh EA. Eco-Epidemiology of Cutaneous Leishmaniasis in Golestan Province, Northeastern Iran: A Systematic Review. Jorjani Biomedicine Journal. 2020;8:60–78.

68. Kassiri H, Naddaf SR, Javadian EA, Mohebali M. First Report on Isolation and Characterization of Leishmania major from Meriones hurrianae (Rodentia: Gerbillidae) of A Rural Cutaneous leishmaniasis Focus in South-Eastern Iran. Iran Red Crescent Med J. 2013;15(9):789-

69. Bellali H, Hchaichi A, Talmoudi K, Harizi C, Chahed M. Effect of climate change on vector-borne diseases: Emerging and increasing incidence of zoonotic cutaneous leishmaniasis in Central Tunisia. Revue d’Épidémiologie et de Santé Publique. 2018;66:S337.

70. Orshan L, Elbaz S, Ben-Ari Y, Akad F, Afik O, Ben-Avi I, et al. Distribution and Dispersal of Phlebotomus papatasi (Diptera: Psychodidae) in a Zoonotic Cutaneous Leishmaniasis Focus, the Northern Negev, Israel. PLoS Negl Trop Dis. 2016;10(7):e0004819.

71. Azarmi S, Zahraei-Ramazani A, Mohebali M, Rassi Y, Akhavan AA, Azarm A, et al. PCR Positivity of Gerbils and Their Ectoparasites for Leishmania Spp. in a Hyperendemic Focus of Zoonotic Cutaneous Leishmaniasis in Central Iran. J Arthropod Borne Dis. 2022;16(2):124–35.

72. Shiravand B, Tafti AAD, Hanafi-Bojd AA, Almodaresi SA, Mirzaei M, Abai MR. Modeling spatial risk of zoonotic cutaneous leishmaniasis in Central Iran. Acta tropica. 2018;185:327–35.

73. Shahabi S, Azizi K, Asgari Q, Sarkari B. Calomyscid Rodents (Rodentia: Calomyscidae) as a Potential Reservoir of Zoonotic Cutaneous Leishmaniasis in a Mountainous Residential Area in the Plateau of Iran: Inferring from Molecular Data of kDNA and ITS2 Genes of Leishmania Major. Journal of Tropical Medicine. 2023;2023.

74. Kassiri H, Sharififard M. A biosystematic and morphometric investigation of the characters of rodents (Mammalia: Rodentia) as reservoir hosts for zoonotic cutaneous leishmaniasis in an endemic focus of Sistan-Baluchistan Province, Iran. Archives of Clinical Infectious Diseases. 2015;10(1).

75. Mirzaei A, Rouhani S, Kazerooni P, Farahmand M, Parvizi P. Molecular detection and conventional identification of leishmania species in reservoir hosts of zoonotic cutaneous leishmaniasis in fars province, South of iran. Iran J Parasitol. 2013;8(2):280–8.

76. Ghafari SM, Ebadatgar V, Mohammadi S, Ebrahimi S, Bordbar A, Parvizi P. Morphologic, Morphometric and Molecular Comparison of Two Sister Species of Rodents as Potential Reservoir Hosts of Zoonotic Cutaneous Leishmaniasis in the Southwest of Iran. Journal of Medical Microbiology and Infectious Diseases. 2019;7(3):79–84.

77. Fatemi M, Yaghoobi-Ershadi MR, Mohebali M, Saeidi Z, Veysi A, Gholampour F, et al. The potential role of humans in the transmission cycle of Leishmania major (Kinetoplastida: Trypanosomatidae), the causative agent of the Old World zoonotic cutaneous leishmaniasis. Journal of Medical Entomology. 2018;55(6):1588–93.

78. Zahirnia AH, Bordbar A, Ebrahimi S, Spotin A, Mohammadi S, Ghafari SM, et al. Predominance of Leishmania major and rare occurrence of Leishmania tropica with haplotype variability at the center of Iran. Braz J Infect Dis. 2018;22(4):278-

79. Jafari R, Najafzadeh N, Sedaghat MM, Parvizi P. Molecular characterization of sandflies and Leishmania detection in main vector of zoonotic cutaneous leishmaniasis in Abarkouh district of Yazd province, Iran. Asian Pacific journal of tropical medicine. 2013;6(10):792–7.

80. Rasi Y, Mohammadi Azni S, Oshaghi MA, Yaghoubi Ershadi M, Mohebali M, Abaei M, et al. Study on Sand Flies as a Vector(s) of Cutaneous Leishmaniasis by Nested PCR in Rural Areas of Damghan District, Semnan Province. Avicenna Journal of Clinical Medicine. 2012;18(4):47–52.

81. Absavaran A, Mohebali M, Moin-Vaziri V, Zahraei-Ramazani A, Akhavan AA, Rafizadeh S, et al. First Report of Natural Infection of Phlebotomus mongolensis to Leishmania major and Leishmania turanica in the Endemic Foci of Zoonotic Cutaneous Leishmaniasis in Iran. Journal of Arthropod-Borne Diseases. 2022;16(4):315–24.

82. Parvizi P, Alaeenovin E, Kazerooni PA, Ready PD. Low diversity of Leishmania parasites in sandflies and the absence of the great gerbil in foci of zoonotic cutaneous leishmaniasis in Fars province, southern Iran. Transactions of the Royal Society of Tropical Medicine and Hygiene. 2013;107(6):356–62.

83. Gholamian-Shahabad MR, Azizi K, Asgari Q, Kalantari M, Moemenbellah-Fard MD. Sandflies species composition, activity, and natural infection with Leishmania, parasite identity in lesion isolates of cutaneous leishmaniasis, central Iran. Journal of Parasitic Diseases. 2018;42:252 – 8.

84. Kassiri H, Najafi S. Bioecology of sandflies (Diptera: Psychodidae: Phlebotominae) in Khorramshahr County, the endemic focus of zoonotic cutaneous leishmaniasis in Khuzestan Province, Iran (2017–2018). International Archives of Health Sciences. 2023;10(1):25–30.

85. Azizi K, Fakoorziba MR, Jalali M, Moemenbellah-Fard MD. First molecular detection of Leishmania major within naturally infected Phlebotomus salehi from a zoonotic cutaneous leishmaniasis focus in southern Iran. Trop Biomed. 2012;29(1):1–8.

86. Cherif K, Boudrissa A, Cherif MH, Harrat Z. [A social program for the control of zoonotic cutaneous leishmaniasis in M’Sila, Algeria]. Sante Publique. 2012;24(6):511–22.

87. Bouderoua K, Benmahdi T, Ammam A. The effect of introducing Mustela nivalis to control the reservoirs of Zoonotic Cutaneous Leishmaniasis at Ain Skhouna in Algeria. Advances in Environmental Biology. 2017;11(3):58–68.

88. Fellahi A, Djirar N, Cherief A, Boudrissa A, Eddaikra N. Zoonotic cutaneous leishmaniasis and leishmania infection among meriones shawi population in setif province, Algeria. Biodiversitas. 2021;22:2547 – 54.

89. Chaouch M, Chaabane A, Ayari C, Ben Othman S, Sereno D, Chemkhi J, et al. Investigation of natural infection of Phlebotomine (Diptera: Psychodidae) by Leishmania in Tunisian endemic regions. Parasite Epidemiology and Control. 2021;14:e00212.

90. Echchakery M, Boussaa S, Kahime K, Boumezzough A. Epidemiological role of a rodent in Morocco: Case of cutaneous leishmaniasis. Asian Pacific Journal of Tropical Disease. 2015;5(8):589–94.

91. Echchakery M, Boussaa S, Ouanaimi F, Boumezzough A. The spatio-temporal distribution of rodent species, potential reservoir hosts of zoonotic cutaneous leishmaniasis in Morocco. 2017.

92. Kahime K, Boussaa S, Idrissi AL-E, Nhammi H, Boumezzough A. Epidemiological study on acute cutaneous leishmaniasis in Morocco. Journal of Acute Disease. 2016;5(1):41–5.

93. Ouanaimi F, Boussaa S, Boumezzough A. Phlebotomine sand flies (Diptera: Psychodidae) of Morocco: results of an entomological survey along three transects from northern to southern country. Asian Pacific Journal of Tropical Disease. 2015;5(4):299–306.

94. Saleh AM, Labib A, Abdel-Fattah MS, Al-Attar MB, Morsy TA. SAND-FLY PHLEBOTOMUS PAPATASI (PHLEBOTOMINAE): A GENERAL REVIEW WITH SPECIAL REFERENCE TO ZOONOTIC CUTANEOUS LEISHMANIASIS IN EGYPT. J Egypt Soc Parasitol. 2015;45(3):525–44.

95. Sawalha S, Al-Jawabreh A, Hjaija D, Ereqat S, Nasereddin A, Al-Jawabreh H, et al. Effectiveness of insecticide thermal fogging in hyrax dens in the control of leishmaniasis vectors in rural Palestine: A prospective study. PLoS Negl Trop Dis. 2022;16(9):e0010628.

96. Khan K, Khan NH, Wahid S. SYSTEMATIC REVIEW OF LEISHMANIASIS IN PAKISTAN: EVALUATING SPATIAL DISTRIBUTION AND RISK FACTORS. J Parasitol. 2021;107(4):630–8.

97. Tomás-Pérez M, Khaldi M, Riera C, Mozo-León D, Ribas A, Hide M, et al. First report of natural infection in hedgehogs with Leishmania major, a possible reservoir of zoonotic cutaneous leishmaniasis in Algeria. Acta tropica. 2014;135:44–9.

98. Tabasi M, Alesheikh AA. Spatiotemporal Variability of Zoonotic Cutaneous Leishmaniasis Based on Sociodemographic Heterogeneity. The Case of Northeastern Iran, 2011-2016. Jpn J Infect Dis. 2021;74(1):7–16.

99. Babaie E, Alesheikh AA, Tabasi M. Spatial modeling of zoonotic cutaneous leishmaniasis with regard to potential environmental factors using ANFIS and PCA-ANFIS methods. Acta Trop. 2022;228:106296.

100. Kalteh EA, Sofizadeh A, Yapng Gharavi AH, Ozbaki GM, Kamalinia HR, Bagheri A, et al. Effect of wild rodents control in reduction of zoonotic cutaneous leishmaniasis in Golestan province, north of Iran (2016). Journal of Gorgan University of Medical Sciences. 2019;21(1):94–100.

101. Jalali H, Enayati AA, Fakhar M, Motevalli-Haghi F, Yazdani Charati J, Dehghan O, et al. Reemergence of zoonotic cutaneous leishmaniasis in an endemic focus, northeastern Iran. Parasite Epidemiology and Control. 2021;13:e00206.

102. Tohidinik HR, Mohebali M, Mansournia MA, Niakan Kalhori SR, Ali-Akbarpour M, Yazdani K. Forecasting zoonotic cutaneous leishmaniasis using meteorological factors in eastern Fars province, Iran: a SARIMA analysis. Trop Med Int Health. 2018;23(8):860–9.

103. Riyad M, Chiheb S, Soussi-Abdallaoui M. Cutaneous leishmaniasis caused by Leishmania major in Morocco: Still a topical question; [Leishmaniose cutanée due à Leishmania major au Maroc: Une question encore d’actualité]. Eastern Mediterranean Health Journal. 2013;19:495 – 501.

104. Hajjaran H, Saberi R, Borjian A, Fakhar M, Hosseini SA, Ghodrati S, Mohebali M. The Geographical Distribution of Human Cutaneous and Visceral Leishmania Species Identified by Molecular Methods in Iran: A Systematic Review With Meta-Analysis. Front Public Health. 2021 Jun 25;9:661674. doi: 10.3389/fpubh.2021.661674. PMID: 34249836; PMCID: PMC8267797.

105. Karmaoui A, Sereno D, El Jaafari S, Hajji L. Seasonal Patterns of Zoonotic Cutaneous Leishmaniasis Caused by L. major and Transmitted by Phlebotomus papatasi in the North Africa Region, a Systematic Review and a Meta-Analysis. Microorganisms. 2022;10(12).

106. Rezai A, Moghaddas E, Bagherpor MR, Naseri A, Shamsian SA. Identification of Leishmania Species for Cutaneous leishmaniasis in Gonabad, Bardaskan and Kashmar, Central Khorasan, 2015. Jundishapur J Microbiol. 2017;10(4):e44469.

107. Masoumeh A, Kourosh A, Mohsen K, Hossein MM, Qasem A, Djaefar M-FM, et al. Laboratory based diagnosis of leishmaniasis in rodents as the reservoir hosts in southern Iran, 2012. Asian Pacific Journal of Tropical Biomedicine. 2014;4:S575–S80.

108. Asl FG, Mohebali M, Jafari R, Akhavan AA, Shirzadi MR, Zarei Z, et al. Leishmania spp. infection in Rhombomys opimus and Meriones libycus as main reservoirs of zoonotic cutaneous leishmaniasis in central parts of Iran: Progress and implications in health policy. Acta Trop. 2022;226:106267.

109. Saberi S, Hejazi SH, Jafari R, Bahadoran M, Akbari M, Soleymanifard S, et al. The Cutaneous Leishmaniasis Reservoirs in Northern Baraan Region of Isfahan, Iran. Journal of Isfahan Medical School. 2013;31(253):1497–507

110. Ghaffari D, Hakimi Parizi M, Yaghoobi Ershadi MR, Sharifi I, Akhavan AA. A survey of reservoir hosts in two foci of cutaneous leishmaniasis in Kerman province, southeast of Iran. J Parasit Dis. 2014;38(3):245–9.

111. Nateghpour M, Akhavan AA, Hanafi-Bojd AA, Telmadarraiy Z, Ayazian Mavi S, Hosseini-Vasoukolaei N, et al. Wild rodents and their ectoparasites in Baluchistan area, southeast of Iran. Trop Biomed. 2013;30(1):72–7.

112. Bordbar A, Parvizi P. High density of Leishmania major and rarity of other mammals’ Leishmania in zoonotic cutaneous leishmaniasis foci, Iran. Trop Med Int Health. 2014;19(3):355–63.

113. Najafzadeh N, Sedaghat MM, Sultan SS, Spotin A, Zamani A, Taslimian R, et al. The existence of only one haplotype of Leishmania major in the main and potential reservoir hosts of zoonotic cutaneous leishmaniasis using different molecular markers in a focal area in Iran. Revista da Sociedade Brasileira de Medicina Tropical. 2014;47:599–606.

114. Sofizadeh A, Hanafi-Bojd AA, Shoraka HR. Modeling spatial distribution of Rhombomys opimus as the main reservoir host of zoonotic cutaneous leishmaniasis in northeastern Iran. J Vector Borne Dis. 2018;55(4):297–304.

115. Doroodgar A, Sadr F, Razavi MR, Doroodgar M, Asmar M, Doroodgar M. A new focus of zoonotic cutaneous leishmaniasis in Isfahan Province, Central Iran. Asian Pacific Journal of Tropical Disease. 2015;5:S54–S8.

116. Yaghoobi-Ershadi MR, Marvi-Moghadam N, Jafari R, Akhavan AA, Solimani H, Zahrai-Ramazani AR, et al. Some Epidemiological Aspects of Cutaneous Leishmaniasis in a New Focus, Central Iran. Dermatol Res Pract. 2015;2015:286408.

117. Azizi K, Parvinjahromi H, Moemenbellah-Fard MD, Sarkari B, Fakoorziba MR. Faunal Distribution and Seasonal Bio-Ecology of Naturally Infected Sand Flies in a New Endemic Zoonotic Cutaneous Leishmaniasis Focus of Southern Iran. J Arthropod Borne Dis. 2016;10(4):560–8.

118. Parvizi P, Akhoundi M, Mirzaei H. Distribution, fauna and seasonal variation of sandflies, simultaneous detection of nuclear internal transcribed spacer ribosomal DNA gene of Leishmania major in Rhombomys opimus and Phlebotomus papatasi, in Natanz district in central part of Iran. Iran Biomed J. 2012;16(2):113–20.

119. Mohammadi-Azni S, Kalantari M, Pourmohammadi B. Molecular Detection of Leishmania Infection in Phlebotomine Sand Flies from an Endemic Focus of Zoonotic Cutaneous Leishmaniasis in Iran. Journal of Arthropod-Borne Diseases. 2022;16(3):233–42.

120. Mohammadi S, Vazirianzadeh B, Fotouhi-Ardakani R, Alaee Novin E, Amirkhani A, Samii J, et al. Tatera indica (Rodentia: Muridae) as the Prior Concern and the Main Reservoir Host of Zoonotic Cutaneous Leishmaniasis on the Border of Iran and Iraq. Jundishapur J Microbiol. 2017;10(1):e42452.

121. Mazaherifar S, Solhjoo K, Abdoli A. Outbreak of cutaneous leishmaniasis before and during the COVID-19 pandemic in Jahrom, an endemic region in the southwest of Iran. Emerg Microbes Infect. 2022;11(1):2218–21.

122. Sofizadeh A, Akbarzadeh K, Allah Kalteh E, Karimi F. Relationship Between the Distribution and Biodiversity of Sand Flies (Diptera: Psychodidae) With the Incidence of Zoonotic Cutaneous Leishmaniasis in Endemic Foci of Golestan Province, Iran. J Med Entomol. 2020;57(6):1768–74.

123. Pourmohammadi B, Mohammadi-Azni S, Kalantari M. Natural infection of Nesokia indica with Leishmania major and Leishmania infantum parasites in Damghan city, Northern Iran. Acta Trop. 2017;170:134–9.

